# Paired wastewater and clinical genomics across metropolitan and hospital catchments reveals SARS-CoV-2 relevant mutations

**DOI:** 10.64898/2026.03.31.26346553

**Authors:** Paula Ruiz-Rodriguez, Alejandro Sanz-Carbonell, Alba Pérez-Cataluña, Pablo Cano-Jiménez, Lidia Ruiz-Roldán, Regina Alandes, Carlos Valiente-Mullor, Concepción Gimeno Cardona, Iñaki Comas, Gloria Sánchez, Fernando González-Candelas, Mireia Coscolla

**Affiliations:** Institute for Integrative Systems Biology (I2SysBio), Spanish National Research Council (CSIC) - University of Valencia, FISABIO Joint Research Unit “Infection and Public Health”, Paterna, Spain; Environmental Virology and Food Safety Lab (VISAFELab), Institute of Agrochemistry and Food Technology, IATA-CSIC, Paterna, Spain; Instituto de Biomedicina de Valencia, IBV-CSIC, Valencia, Spain; Servicio de Microbiología, Consorcio Hospital General Universitario, Valencia, Spain; Facultad de Medicina, Departamento de Microbiología, Universidad de Valencia, Valencia, Spain; CIBERESP, Consorcio de Investigación Biomédica en Red de Epidemiología y Salud Pública, Madrid, Spain

**Keywords:** Wastewater, SARS-CoV-2, GWAS, Omicron, hospitalisation

## Abstract

Wastewater (WW) genomics can track SARS-CoV-2 circulation beyond clinical testing, but its ability to reflect clinical diversity and capture severity-linked mutations remains unclear. Here, we integrated 845 clinical genomes and 22 wastewater genomes from Valencia, Spain, across matched metropolitan and hospital catchments. We compared matched WW and clinical sequencing for lineage and mutation surveillance at two levels: metropolitan and hospital. Then, we tested WW sensitivity to detect mutations statistically associated with hospitalization status in regional (n = 4,843), national (n = 10,052) and supranational (n = 39,099) clinical datasets. WW surveillance captured the dominant Omicron background when collapsing lineages into parental lineages constellation but had limited sensitivity for fine-scale sublineage diversity. Performance was strongly catchment dependent: metropolitan wastewater best represented broader community circulation, whereas hospital wastewater was noisier but detected KP.3 months before its appearance in routine metropolitan clinical surveillance. Across clinical datasets, hospitalisation-associated substitutions showed limited reproducibility, although the national and supranational analyses converged on receptor-binding-domain substitutions D405N, K417N and R408S. Networks showed coupling between G252V in NTD with those RBD substitutions involved in immune escape and receptor engagement. Finally, integrating regional to supranational GWAS with interaction networks and wastewater detection prioritised mutations supported by at least two independent association layers, that includes mutations in the Spike, especially in RBD, and the wastewater-exclusive candidate S:V445P, which was missed by contemporaneous clinical sequencing. Overall, WW genomics preferentially recovers the common mutational backbone of SARS-CoV-2 circulation and can highlight important changes missed by clinical sampling, making it a complementary tool for real-time prioritisation of viral evolutionary change.

## Introduction

As SARS-CoV-2 surveillance has shifted from emergency response to longer-term public health monitoring, wastewater-based surveillance has emerged as a valuable complement to clinical surveillance because it captures population-level viral circulation independently of individual testing. This role becomes especially important when surveillance must resolve epidemiologically meaningful change within a lineage background that already dominates transmission. This is the challenge posed with the Omicron-dominating phase, when diversification among closely related sublineages allowed mutations with potential effects on antigenicity, immune escape and transmission fitness to spread without clear lineage replacement outside the Omicron background^1–3^. As routine clinical ascertainment declined, reduced testing and incomplete sequencing could distort both the apparent prevalence of circulating sublineages and the mutational spectrum available for downstream analysis^4,5^. Under these conditions, lineage labels alone were often insufficient, and meaningful surveillance increasingly required tracking the specific mutations accumulating within circulating viral subpopulations^1,2^. These same constraints also complicated attempts to relate viral genotype to clinical outcome, because the sequences available for analysis reflected not only viral circulation, but also testing strategies and sample representativeness^4,6^. A complementary population-level view of viral circulation that was less dependent on individual testing therefore became increasingly valuable^7^.

Wastewater-based surveillance can capture community transmission independently of individual testing and can support public health decision-making when clinical sampling becomes sparse^7,8^. Early work showed that wastewater RNA trajectories can track and even anticipate traditional indicators, including hospital admissions, highlighting its value as a population-level sensor^9–11^. Extending wastewater monitoring from quantification to genomic characterisation has further demonstrated that wastewater sequencing can detect variant turnover and “cryptic” transmission that is not represented in clinical genomic datasets^12–16^. Importantly, SARS-CoV-2 shed into sewage integrates contributions from symptomatic and asymptomatic infections, generating a population-level composite of recent viral circulation^7,8,17^. This broader sampling in wastewater is especially valuable during later Omicron circulation, when declining diagnostic testing reduced clinical ascertainment while ongoing transmission and within-Omicron diversification continued^2,4^.

Environmental sequencing, however, is not simply clinical genomics applied to a larger sample. Wastewater RNA is fragmented, genome coverage is heterogeneous and each sample contains mixtures of viral templates derived from multiple infected individuals^12,18,19^. A substitution can be detected without full haplotype reconstruction, whereas a true circulating mutation may be missed because of low abundance, uneven coverage or stochastic loss in mixed templates^12,18^. Detection probability is therefore intrinsically linked to both prevalence in the contributing population and local genome recoverability. In mixed and unevenly covered wastewater samples, this should favour more consistent recovery of common background substitutions than of rarer mutations, which are more likely to be missed and may include the earliest emerging changes^18,20,21^. These analytical constraints motivate benchmarking against matched clinical data^12,19^. They also make catchment context central to interpretation. A metropolitan wastewater treatment plant integrates shedding from a large and heterogeneous urban population and tends to smooth local fluctuations through mixing and sewer network transport^22,23^. In contrast, sampling from a local sewer collector that serves a specific institution or facility represents a smaller and more structured source population, which may introduce different forms of stochasticity and representativeness^23–25^. Wastewater performance cannot therefore be assumed to be uniform across surveillance systems, and lineage- and mutation-level concordance must be evaluated in relation to the specific catchment being sampled.

In the Omicron era, functionally important changes often accumulate in a limited set of recurrent mutational hotspots, with the same residues arising independently in multiple backgrounds^2,3,26^. The phenotypic and epidemiological interpretation of a given substitution can therefore depend on its surrounding mutation set and on the immunity landscape in which it circulates^3,27^. In this setting, we have used a combined analytical strategy that pairs marginal association testing with regularised interaction-network modelling. The first captures population-level statistical enrichment, whereas the second is intended to expose conditional structure that may be obscured when substitutions are analysed one at a time. For wastewater, where direct haplotype reconstruction is often incomplete, this approach is particularly useful because it enables prioritisation of environmental signals without over-interpreting fragmented mixed-template data as fully resolved genomes^18,19^.

In this context, and aligned with WHO guidance that emphasises integrating wastewater and clinical evidence streams, our study combined catchment-matched clinical and wastewater sequencing in Valencia, Spain during Omicron expansion across two epidemiologically distinct datasets, a metropolitan and a hospital, and used this framework to interpret wastewater-detectable lineages and mutations in relation to clinical outcomes^28^. The study integrates two complementary environmental systems, a metropolitan wastewater treatment plant serving a large urban catchment and a hospital sewer collector representing a healthcare-associated source population, together with matched clinical genomes from hospitalised and non-hospitalised infections. It further extends hospitalisation association analyses to regional, national and supranational clinical cohorts, allowing environmental detections to be evaluated against clinical evidence across epidemiological scales. This design supports three linked levels of analysis: comparison of wastewater and clinical lineage composition across distinct catchments, coverage-aware evaluation of mutation recovery in environmental samples, and prioritisation of wastewater-detectable mutations using convergence between univariate association and interaction-aware multivariable models trained on outcome-annotated clinical data.

## Results

### Wastewater sequencing captures dominant lineage dynamics and reveals lineage emergence ahead of clinical surveillance

We assessed how accurately wastewater (WW) sequencing recapitulated the lineage diversity observed in clinical infections in two surveillance settings in Valencia, Spain: a metropolitan catchment and a hospital (Fig. 1A). The metropolitan dataset comprised 11 WW samples and 744 clinical genomes (216 from hospitalised cases and 528 from non-hospitalised cases) collected between 9 July and 8 December 2023 (Fig. 1B; Supplementary Fig. S1, panel A). Hospitalised and non-hospitalised clinical samples shared 16 of 21 lineage constellations (Jaccard index = 0.762) and mean absolute error (MAE) between the two clinical series was only 2.31 percentage points, significantly lower than the MAE between WW and pooled clinical data (4.09 percentage points; Welch’s *t* = 3.19, *P* = 0.005; Cohen’s *d* = 1.36). Consequently, we used the pooled clinical series as the primary comparator for metropolitan analyses against WW (Supplementary Table SA and Supplementary Table SB). At the broad clade level, metropolitan WW shared 7 of 14 clades with pooled clinical surveillance (Jaccard index = 0.50; Supplementary Table SA), indicating moderate qualitative overlap. At the finer collapsed lineage-constellation level, defined using the remapping scheme in Supplementary Table S3, WW recovered only 6 of the 21 constellations detected clinically (Jaccard index = 0.286; sensitivity = 28.6%; Supplementary Table SA), but all WW-detected constellations were also present in clinical surveillance (positive predictive value = 100%; Fig. 1D; Supplementary Table SA). Thus, metropolitan WW captured only a subset of circulating lineages, but that subset corresponded to the dominant lineage families. Consistent with this, abundance-based concordance improved at the constellation level relative to clade level, with mean Jensen-Shannon divergence (JSD) decreasing from 0.613 to 0.277 and mean Bray-Curtis dissimilarity from 0.960 to 0.459 (Supplementary Table SA). Constellation abundances were also significantly correlated between WW and clinical surveillance (Spearman’s ρ = 0.564, P < 0.001), with a mean MAE of 4.09 percentage points across matched windows (Supplementary Table SA). Window-level MAE values ranged from 1.74 to 6.35 percentage points, with the lowest errors in September and higher values during periods of lineage turnover (Supplementary Fig. S2; Supplementary Table SB). Temporal analyses showed that similarity between WW and clinical lineage profiles was strongly affected by parental lineage reassignment (Supplementary Fig. S3). When reassignment was applied (Supplementary Fig. S3, panel A), similarity between WW and both hospitalised and non-hospitalised cases ranged from 0.25 to 1.00. Without reassignment (Supplementary Fig. S3, panel B), concordance was substantially lower and more variable. Consistent with a community-integrated signal, metropolitan WW profiles were markedly less volatile across successive windows than metropolitan clinical profiles, as also evident from the smoother temporal composition in Fig. 1F and Supplementary Fig. S4, panel A.

**Figure 1.**
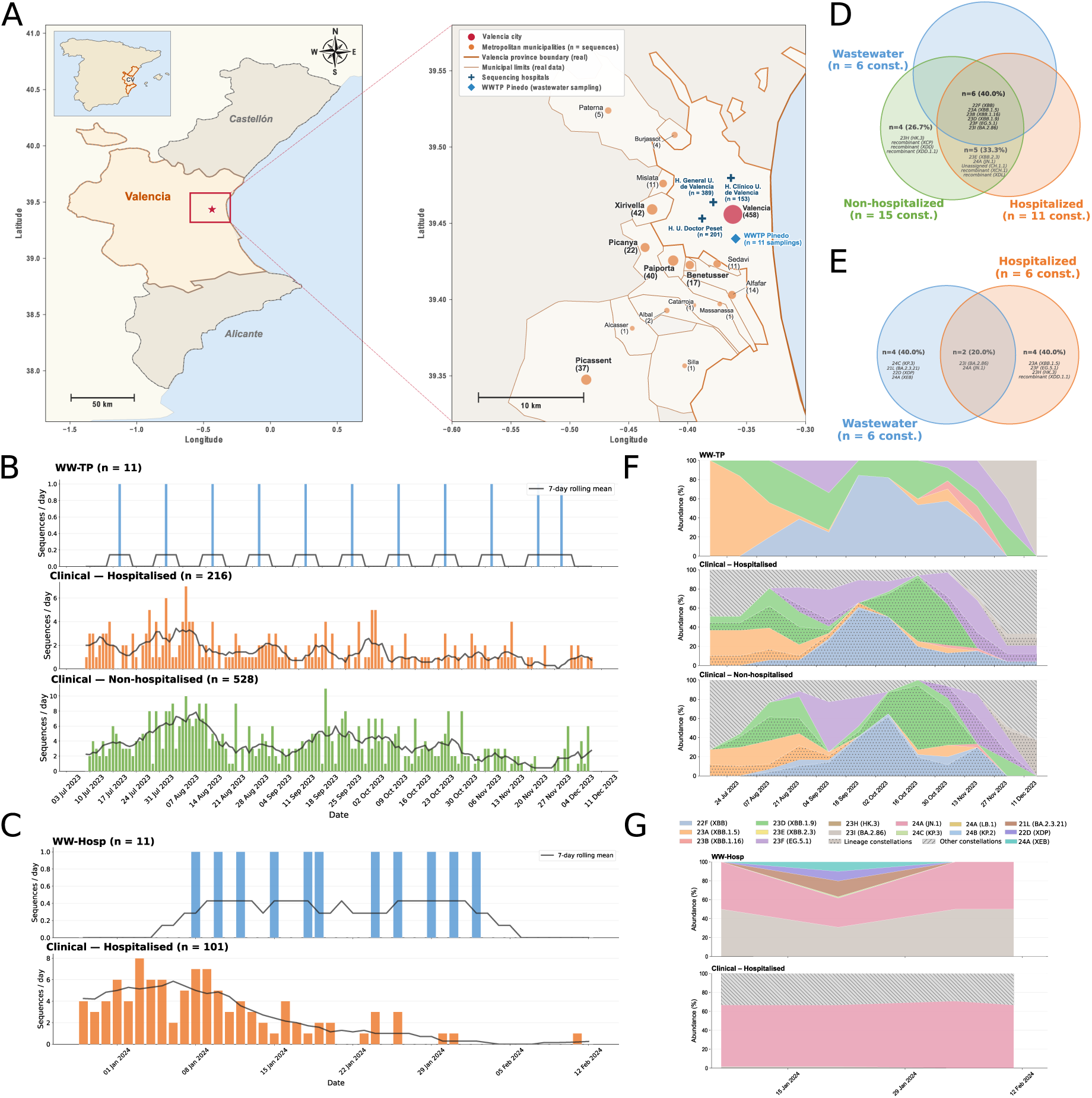
Sampling design and SARS-CoV-2 lineage dynamics in matched clinical and wastewater surveillance datasets from the Valencia metropolitan area and Hospital General Universitario de Valencia. **(A)** Geographic context and surveillance catchments. Left, location of Valencia Province within Spain and of the metropolitan area analysed. Right, municipalities included in the metropolitan catchment drain to the Pinedo wastewater treatment plant (WW-TP). Circles denote municipalities of residence of sequenced clinical cases and are scaled to the number of genomes, with counts shown next to labels. Blue crosses indicate sequencing hospitals, with the corresponding number of genomes shown beside each hospital, and the blue diamond marks the WW-TP sampling point. The hospital dataset comprised only hospitalised cases and wastewater collected from the hospital sewer collector (WW-Hosp). **(B,C)** Temporal distribution of sequenced clinical genomes and wastewater samples in the metropolitan dataset **(B)** and hospital dataset **(C)**. Bars show the number of genomes or wastewater samples collected per day, and black lines indicate the 7-day rolling mean. **(D,E)** Venn diagrams showing overlap in lineage constellations among wastewater and clinical datasets for the metropolitan comparison **(D)** and hospital comparison **(E)**. Counts and percentages refer to the total number of distinct constellations represented in each diagram. **(F,G)** Temporal changes in clade and lineage-constellation composition across matched sampling windows in the metropolitan dataset **(F)** and hospital dataset (**G**), shown as stacked area plots of relative abundance. Nextclade clades are annotated with representative Pango lineages in parentheses. Areas with dotted fills denote lineage constellations comprising multiple closely related Pango lineages, whereas diagonally hatched grey regions indicate constellations grouped as Other constellations not present in WW.

Temporal concordance in the metropolitan dataset depended strongly on parental lineage reassignment. After collapsing clinical sublineages to their parental constellations, Jaccard similarity between WW and both hospitalised and non-hospitalised cases ranged from 0.25 to 1.00, with perfect agreement in July-August and October and transient declines in mid-September and late November (Supplementary Fig. S3, panel A). Without parental reassignment, concordance was lower and more variable, and WW versus hospitalised cases fell to J = 0.00 in September (Supplementary Fig. S3, panel B). Overall, collapsing sublineages to their parental constellations increased metropolitan WW-clinical Jaccard similarity from 0.286 to 0.667, indicating that most discordance arose from fine-scale sublineage fragmentation and recombinant lineages rather than from disagreement in the dominant Omicron backgrounds. The effect of the remapping strategy on quantitative error is further illustrated in Supplementary Fig. S2, and the corresponding lineage-collapsing rules are detailed in Supplementary Table S3. Metropolitan WW profiles were also less volatile across successive windows than the corresponding hospitalised and non-hospitalised clinical profiles, consistent with the smoother temporal dynamics visible in Fig. 1F and Supplementary Fig. S4, panel A.

We observed the same overall pattern in the hospital dataset, which comprised 101 clinical genomes and 11 WW sampling dates collected between 29 December 2023 and 12 February 2024 (Fig. 1C; Supplementary Fig. S1, panel B), but with poorer qualitative overlap and greater quantitative error. At the clade level, the overlap between WW and clinical surveillance at the hospital was weak, with only two shared clades (Jaccard index = 0.222), and relative abundances were discordant, including an inverse rank relationship between WW and clinical clade profiles. For quantitative lineage-constellation comparisons, the mapped WW repertoire was restricted to three standard constellations, of which only BA.2.86 and JN.1 were shared with clinical surveillance; under this representation, concordance remained modest (Jaccard index = 0.250; sensitivity = 28.6%; positive predictive value = 66.7%), although abundance-based agreement was again stronger than at clade level (mean JSD = 0.527; mean Bray–Curtis dissimilarity = 0.653; Spearman’s ρ = 0.360, *P* < 0.001; Fig. 1E,G; Supplementary Table SA). Even so, the quantitative mismatch remained substantially larger than in the metropolitan dataset, with a mean MAE of 11.97 percentage points across matched windows (Supplementary Table SA; Supplementary Table SB). Raw WW deconvolution additionally yielded BA.2.3.21, XDP and XEB without contemporaneous clinical counterparts, which further expanded the hospital WW repertoire and reduced raw presence–absence concordance (Supplementary Fig. S5, panel A).

The distinction between collapsed constellations and complete lineage WW calls was especially informative for emergence signals. In the hospital dataset, raw WW detections comprised six lineage categories overall, but only two were shared with clinical surveillance, yielding a raw Jaccard index of 0.182 and a lower-than-expected overlap under permutation (*P* = 0.049). Among the WW-exclusive calls, KP.3 (24C) was first detected in hospital WW on 12 January 2024 and reappeared on 2 February 2024, while remaining absent from contemporaneous hospital clinical sequencing (Supplementary Fig. S5, panel B). In the extended metropolitan clinical dataset, which spanned June 2023 to August 2024 and extended beyond the July–December 2023 comparison window analysed here, KP.3 was not observed until May 2024, indicating that WW detected this lineage several months before its appearance in routine clinical surveillance (Supplementary Fig. S5, panel C). The same hospital WW series also contained BA.2.3.21, XDP and XEB, indicating that WW can capture transient or weakly represented lineage signals not recovered by matched clinical sequencing.

Together, these results show that WW sequencing provides an incomplete but highly informative representation of circulating SARS-CoV-2 diversity.

### Wastewater sequencing captures common circulating SARS-CoV-2 but misses rare mutations

We next compared individual SARS-CoV-2 mutations between wastewater (WW) and clinical surveillance. Accounting for coverage heterogeneity across wastewater samples (using a coverage-aware, time-window framework that retained only positions reliably sequenced in each WW sample; Supplementary Fig. S6, panels A and B; Supplementary Fig. S7, panels A-D; Supplementary Table S4) substantially improved the estimated recovery of clinically observed mutations. Consensus-based comparisons implicitly penalised WW at positions that were not reliably covered in all samples, thereby underestimating sensitivity. When comparisons were restricted to temporally matched windows and to positions adequately covered (according to the quality thresholds described in the Methods) in each WW sample, sensitivity increased from 0.029 to 0.103 in the metropolitan dataset (3.5-fold; adjusted P = 5.48 × 10⁻²⁵) and from 0.075 to 0.368 in the hospital dataset (4.9-fold; adjusted P = 3.77 × 10⁻²¹), while predictive value (PPV)remained high (Supplementary Fig. S7). Thus, sample-specific coverage masks provide a more accurate benchmark of WW performance against clinical surveillance. Consequently, we focused on using a coverage-aware, time-window framework that retained only positions reliably sequenced in each WW sample (Supplementary Fig. S6; Supplementary Fig. S7; Supplementary Table S4). WW displayed a consistent profile of high positive PPV but low sensitivity in the 1,549 mutations in the metropolitan dataset and 322 in the hospital dataset. In the metropolitan comparison, 151 mutations were shared between WW and clinical surveillance (Jaccard index = 0.097; sensitivity = 0.103, 95% CI = 0.088-0.120; PPV = 0.651, 95% CI = 0.588-0.709), whereas 103 mutations were shared in the hospital comparison (Jaccard index = 0.320; sensitivity = 0.368, 95% CI = 0.314-0.426; PPV = 0.710, 95% CI = 0.632-0.778) (Fig. 2A,B). Most mutations were detected only in clinical samples (1,317 in the metropolitan dataset and 177 in the hospital dataset), whereas WW-only mutations were less frequent (81 and 42 sites), indicating that mutations recovered from WW were often corroborated clinically, but that WW captured only a subset of the mutational diversity observed in patients (Fig. 2A,B; Supplementary Table S4).

**Figure 2.**
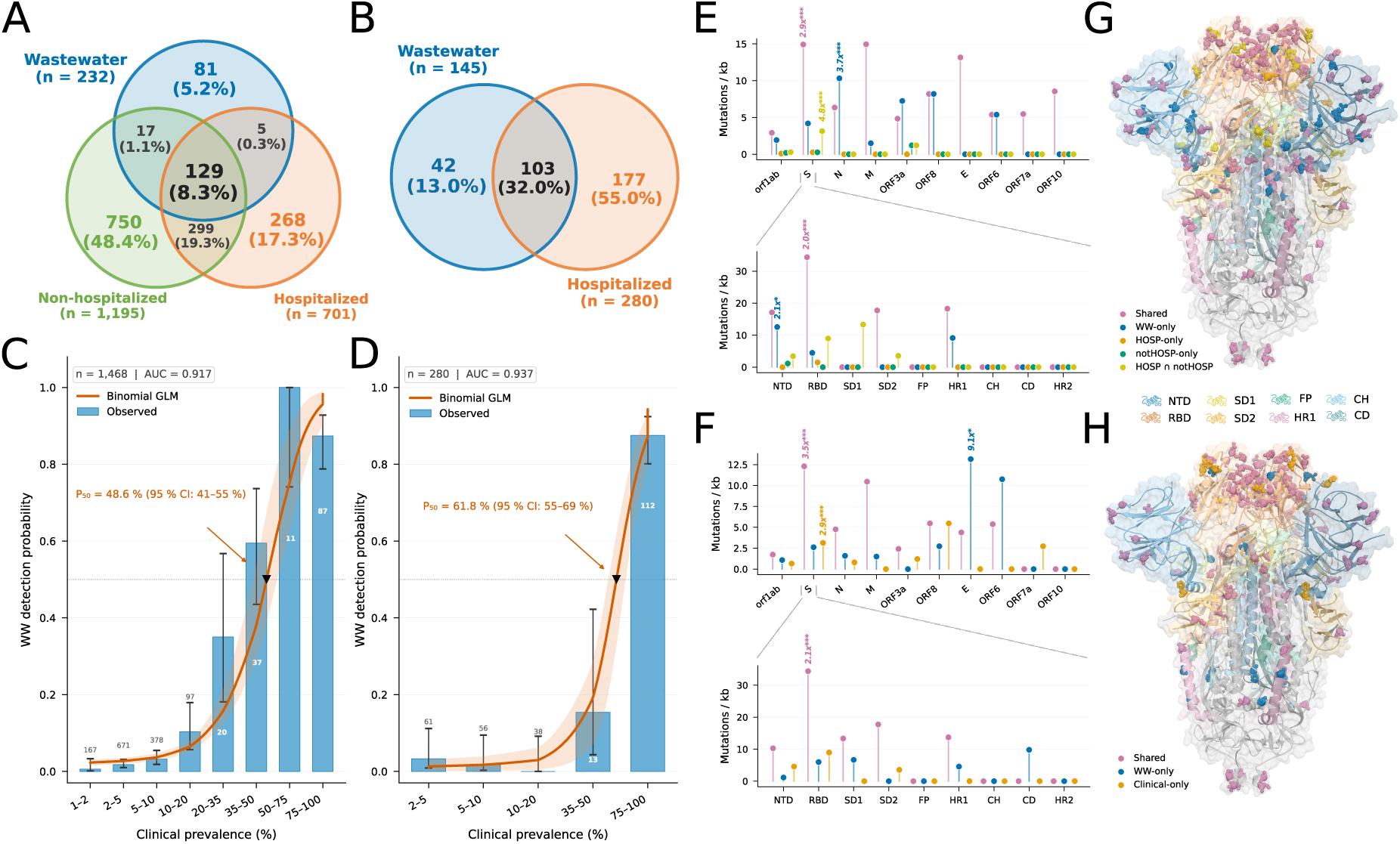
Mutation-level concordance between wastewater and clinical SARS-CoV-2 surveillance in the metropolitan and hospital datasets. **(A,B)** Venn diagrams showing the overlap among distinct SARS-CoV-2 mutational positions detected in wastewater (WW) and clinical samples in the metropolitan dataset (**A**) and the hospital dataset (**B**). Numbers indicate mutational positions, and percentages indicate the proportion of all evaluable positions in each dataset. **(C,D)** WW detection probability as a function of clinical mutation prevalence in the metropolitan dataset (**C**) and hospital dataset (**D**). Blue bars show observed WW detection frequencies for mutations grouped by clinical prevalence; error bars denote Wilson 95% confidence intervals, and numbers indicate the number of mutations in each bin. Orange curves show binomial generalised linear models (GLMs; logit link) fitted by maximum likelihood, with 95% bootstrap confidence bands (shaded; 2,000 resamples). Inverted triangles mark *P*₅₀, the clinical prevalence at which the fitted probability of WW detection reaches 0.5. The corresponding area under the curve (AUC) and *P*₅₀ values are shown in each panel. **(E,F)** Length-normalised mutation densities across SARS-CoV-2 genes (upper panels) and Spike domains (lower panels) in the metropolitan dataset (**E**) and hospital dataset (**F**). Lollipops show mutation density (mutations kb⁻¹) for each gene or Spike domain, coloured by detection category. Selected fold-enrichment values relative to length-proportional null expectations are annotated above lollipops; significance was assessed using one-sided binomial tests with Bonferroni correction (*P* < 0.05, *P* < 0.01, *P* < 0.001). Dashed lines connect the Spike gene to the corresponding domain-level analysis. **(G,H)** Spatial distribution of Spike mutations in the metropolitan dataset (**G**) and hospital dataset (**H**). The prefusion Spike trimer is coloured by structural domain, and residues carrying mutations are shown as spheres coloured according to detection category, as in E and F.

Clinical prevalence was the main determinant of WW detection (Fig. 2C,D). In binomial generalised linear models, clinical prevalence predicted WW detection with excellent discrimination in both settings (AUC = 0.917 in the metropolitan dataset, n = 1,468; AUC = 0.937 in the hospital dataset, n = 280). The fitted curves showed a steep transition in detection probability, with an estimated P₅₀ of 48.6% (bootstrap 95% CI = 41-55%) in the metropolitan dataset and 61.8% (95% CI = 55-69%) in the hospital dataset. Consistently, mutations present in ≤50% of clinical samples were recovered in WW in only 4.7% and 3.0% of cases, respectively, whereas mutations present in >50% of clinical samples were detected in 88.8% and 87.5% of cases. Thus, WW reliably recovered the common mutational backbone of circulating viruses, but seldom captured low-prevalence mutations, including those at the leading edge of emergence (Fig. 2C,D; Spearman’s ρ = 0.800, P = 6.43 × 10⁻³⁰ in the metropolitan dataset; ρ = 0.440, P = 1.04 × 10⁻² in the hospital dataset).

This prevalence dependence was also reflected in temporal concordance across matched windows. In the metropolitan dataset, Jaccard similarity increased from 0.161 in early windows to 0.245 in late windows, sensitivity from 0.166 to 0.256, and PPV from 0.834 to 0.864. The same trend was observed in the hospital dataset, where Jaccard similarity increased from 0.503 to 0.648 and sensitivity from 0.543 to 0.721, whereas PPV remained essentially stable (0.866 to 0.868) (Supplementary Fig. S8). Together, these results indicate that WW concordance improved as mutational diversity narrowed, but remained constrained for rarer variants.

Mutation sharing was not evenly distributed across the genome (Fig. 2E,F; Supplementary Fig. S9). Shared mutations were concentrated mainly in *S* and *M* genes, whereas *orf1ab* contributed many shared sites in absolute terms but was not enriched after normalising for gene length. Wastewater detection of clinically observed mutations was highest for small structural genes, with complete sensitivity for *M* and *E* in the metropolitan dataset and for *M* in the hospital dataset, while *S* and *orf1ab* showed high but incomplete sensitivity in both settings. Accessory genes displayed more variable detection patterns, including lower sensitivity for *orf3a* in the metropolitan dataset and no wastewater detection of *orf7a* in the hospital dataset. WW-only mutations showed a broader and more heterogeneous gene distribution, with additional representation in *N*, *S* and *orf3a* in the metropolitan comparison and in *E* in the hospital comparison (Fig. 2E,F; Supplementary Fig. S9).

Within Spike, shared nucleotide mutations clustered primarily in the N-terminal domain (NTD) and receptor-binding domain (RBD), whereas distal S2 regions carried few recurrent changes (Fig. 2G,H). In the length-normalised domain analysis, the RBD was enriched among shared mutations in Spike, whereas WW-only mutations in Spike showed additional enrichment in the NTD in the metropolitan dataset (Supplementary Fig. S10). When domain distributions were compared across detection categories, however, only the enrichment of WW-only NTD mutations in the metropolitan dataset remained significant after multiple-testing correction (OR = 6.02, adjusted P = 2.47 × 10⁻²), and no Spike-domain enrichment remained significant in the hospital dataset (Supplementary Fig. S10, panel B). Overall, these results show that WW sequencing provides a robust readout of the common mutational backbone of circulating SARS-CoV-2 and can reveal mutations not observed in matched clinical surveillance, but incompletely captures the low-frequency tail of mutational diversity and the earliest stages of mutational emergence (Fig. 2; Supplementary Table S4).

### Hospitalisation-associated amino acid signals are predominantly protective but only partly reproducible across scales

We investigated whether recurrent SARS-CoV-2 amino acid substitutions were associated with hospitalisation outcome across three GWAS datasets: a regional cohort from the Comunitat Valenciana (*n* = 4,843) analysed using univariate regression, and national (Spain, *n* = 10,052) and supranational (*n* = 39,099) datasets, using logistic regression (Fig. 3A-C; Supplementary Table SX). The three hospitalisation datasets also differed in phylogenetic composition and host demographic structure (Supplementary Fig. S11, panels A-D). After removing minor allele frequency mutations, only non-synonymous substitutions were considered, 143, 149 and 160 variants were tested in the regional, national and supranational datasets, respectively (Supplementary Table S6). The number of Bonferroni-significant associations increased with cohort size, from 5/143 (3.5%) in the regional cohort to 8/149 (5.4%) in the national cohort and 35/160 (21.9%) in the supranational cohort, consistent with greater power at larger scale. Significant signals were dominated by substitutions associated with lower odds of hospitalisation: all five regional hits were protective, as were 5/8 national hits and 31/35 supranational hits. The regional cohort yielded five protective associations, nsp10:S33C, S:Q493E, nsp3:T1465I, S:V1104L and S:F456L, whereas the national cohort identified five protective Spike substitutions (S:V83A, S:K417N, S:R408S, S:N501Y and S:Y505H) together with three risk-associated substitutions (S:D405N, nsp6:R252K and S:G252V). The supranational cohort detected four risk-associated substitutions (S:D796Y, S:D405N, S:E484A and S:Q498R), but was again dominated by protective signals, including nsp3:A1892T, N:Q229K, M:A104V and nsp2:A31D. The strongest associations by statistical support were S:Q493E in the regional cohort (log₂(OR) = −0.944, *P* = 9.14 × 10⁻⁹), S:V83A in the national cohort (log₂(OR) = −0.496, *P* = 1.65 × 10⁻²⁵) and S:D796Y in the supranational cohort (log₂(OR) = +0.241, *P* = 1.86 × 10⁻⁷⁰).

**Figure 3.**
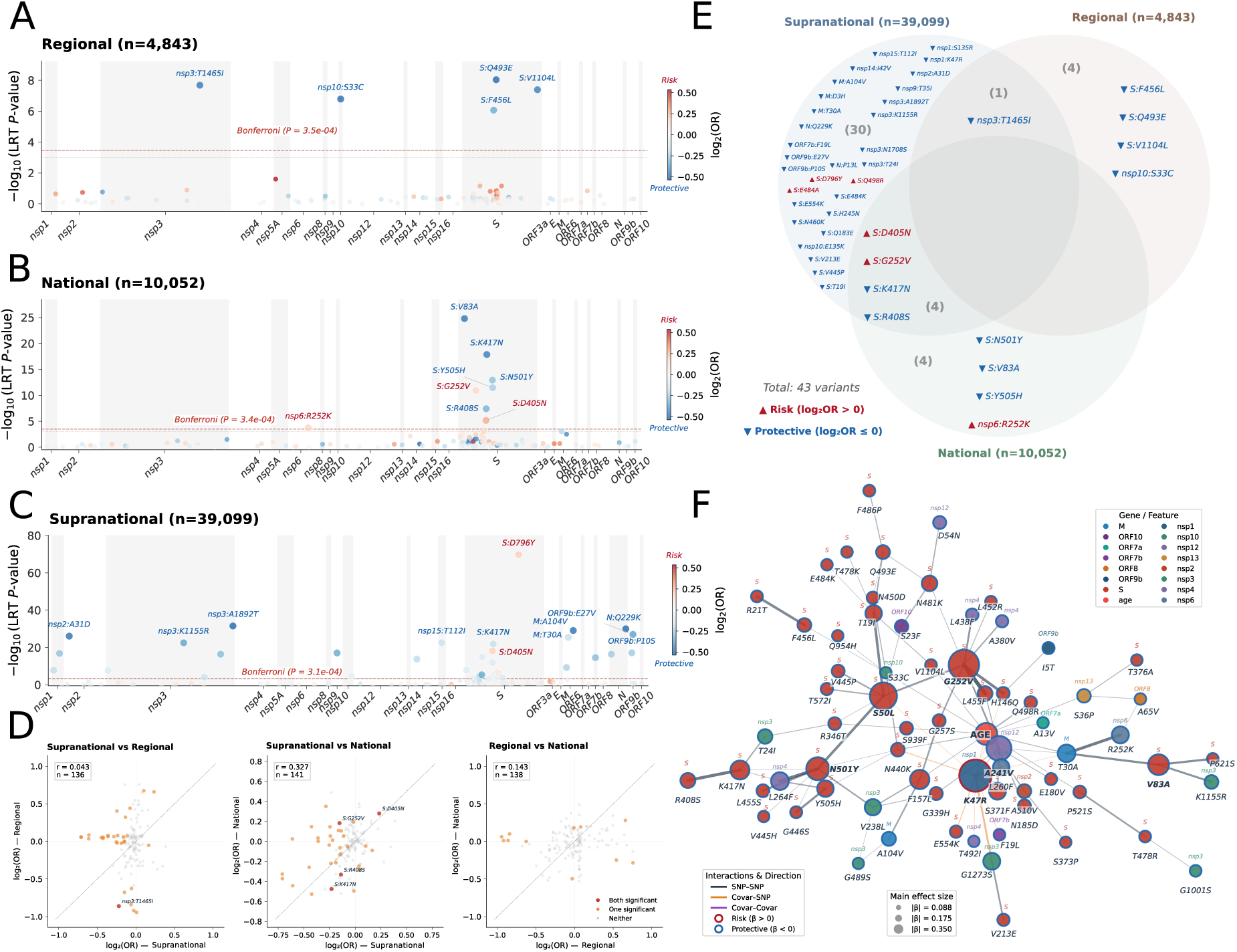
Amino acid substitutions associated with hospitalisation outcome across regional, national and supranational SARS-CoV-2 cohorts. **(A–C)** Manhattan-style plots showing single-variant associations between amino acid substitutions and hospitalisation outcome (hospitalised versus non-hospitalised infections) in the regional cohort (**A**; *n* = 4,843), the national cohort (**B**; *n* = 10,052) and the supranational cohort (**C**; *n* = 39,099). Each point represents one amino acid substitution, positioned along the SARS-CoV-2 genome by gene and amino acid coordinate. The y axis shows statistical support as −log₁₀(*P*) from the likelihood-ratio test, and point colour indicates effect size as log₂(odds ratio, OR), with red denoting increased odds of hospitalisation and blue denoting decreased odds (protective association). Horizontal dashed red lines indicate the Bonferroni-corrected significance threshold (α = 0.05/*m*). Selected Bonferroni-significant substitutions are annotated. **(D)** Pairwise concordance of log₂(OR) estimates for shared variants across cohorts. Each point represents one amino acid substitution detected in both cohorts being compared. The diagonal indicates perfect agreement in effect size between cohorts. Point colour denotes whether the variant was Bonferroni-significant in either cohort, one cohort only, or both cohorts. Pearson’s correlation coefficient (*r*) and the number of shared variants (*n*) are shown in each panel. **(E)** Venn diagram summarising overlap among Bonferroni-significant amino acid substitutions across the three cohorts (total = 43 variants). Variants are labelled within each sector and coloured according to direction of effect: red, risk-associated (log₂(OR) > 0); blue, protective (log₂(OR) < 0). **(F)** In the national level (spanish cohort) the network visualises the glinternet model: nodes represent predictors (amino acid variants and covariates), node size is proportional to the absolute main-effect coefficient (|β|), node fill indicates the gene/covariate, and node outline denotes direction (β > 0, risk-hospitalised; β < 0, protective-non-hospitalised). Edges connect pairs of predictors with non-zero interaction coefficients (edge width scales with interaction strength; edge colour indicates interaction 16 class). Interactions represent model-selected statistical dependencies for binary hospitalization traits.

Cross-cohort reproducibility was limited (Fig. 3E). Across the three GWAS analyses, the union of Bonferroni-significant substitutions comprised 43 variants, of which 30/43 (69.8%) were unique to the supranational cohort, 4/43 (9.3%) were unique to the regional cohort and 4/43 (9.3%) were unique to the national cohort. The remaining 5/43 (11.6%) were significant in exactly two cohorts, and no substitution reached Bonferroni significance in all three. Pairwise overlap tests supported this scale dependence. The supranational and regional cohorts shared only one significant hit, nsp3:T1465I, indicating very limited overlap between both analyses (1.05-fold; Fisher/hypergeometric P = 0.660), whereas the supranational and national cohorts shared four hits, S:D405N, S:G252V, S:K417N and S:R408S, representing 2.52-fold enrichment over expectation (*P* = 0.0497). Among the five substitutions replicated in at least two cohorts, effect directions were concordant for four (80%), comprising three consistently protective substitutions and one consistently risk-associated substitution, whereas S:G252V showed opposite directions in the supranational and national datasets.

Effect-size concordance across all shared tested variants was likewise cohort-dependent (Fig. 3D). Correlation of log₂(OR) estimates was significant only between the supranational and national cohorts (r = 0.327, *P* = 7.38 × 10⁻⁵; bootstrap 95% CI = 0.139-0.494), but not between the supranational and regional cohorts (r = 0.043, *P* = 0.617) or the regional and national cohorts (r = 0.143, *P* = 0.0949). Consistent with this, Cochran’s Q tests identified significant effect heterogeneity for 39/147 shared substitutions (*P* < 0.05), including four of the five replicated hits (S:G252V, nsp3:T1465I, S:K417N and S:R408S), whereas S:D405N showed no evidence of between-cohort heterogeneity (Q-test *P* = 0.527; Supplementary Table S7). Together, these results indicate that amino acid associations with hospitalisation are only partly reproducible across epidemiological scales, despite a small core of recurring Spike signals.

To move beyond marginal associations, we fitted regularised interaction models (GLINTERNET) in the national and supranational cohorts and summarised the selected dependencies as interaction networks (Fig. 3F, Supplementary Fig. S12, Supplementary Table S8). The national λ_min_ model (minimum cross-validation error penalty parameter) contained 69 nodes and 95 interactions and was dominated by viral substitutions in Spike. The largest main effect corresponded to nsp1:K47R (|β| = 0.350, risk-associated), while the strongest pairwise interactions included S:G252V × S:L455F (0.365), S:R408S × S:K417N (0.278) and nsp6:R252K × M:T30A (0.273). Age (14 interactions) and nsp1:K47R (13 interactions) emerged as the principal hubs. In the more parsimonious national λ_1se_ model (one standard error of the minimum cross-validated error), nsp1:K47R remained the dominant main effect (|β| = 0.654), whereas S:G252V became the central interaction hub. The supranational λ_min_ model was smaller, with 19 nodes and 29 interactions, and more strongly structured by host covariates. Sex and nsp1:K47R each participated in 11 interactions, and the strongest terms were sex × nsp1:K47R (0.597), age × sex (0.493) and age × nsp1:K47R (0.454). These signals persisted in the supranational λ_1se_ model, in which sex × nsp1:K47R remained the strongest interaction (0.535). In both cohorts, inclusion of interaction terms improved fit relative to main-effects-only models, and bootstrap selection frequencies indicated greater stability for the supranational than for the national network. Overlap between national and supranational networks was low, with 9/79 shared nodes and 1/122 shared edges at λ_min_, and 3/20 shared nodes but no shared edges at λ_1se_, indicating that interaction structure was largely cohort-specific. Nevertheless, nsp1:K47R emerged consistently across both cohorts and penalty levels. Notably, its marginal association in the supranational GWAS was protective (log₂(OR) = −0.242, *P* = 1.89 × 10⁻⁸), whereas its conditional effect in both interaction models was risk-associated, suggesting that its apparent effect depends on adjustment for higher-order dependencies and host covariates. These network-selected edges should therefore be interpreted as regularisation-derived statistical dependencies rather than direct biological interactions.

### Wastewater surveillance captures hospitalisation-associated mutational signatures

To identify WW-detectable amino acid substitutions with potential relevance to clinical severity outcome, we integrated different analyses of association to hospitalization in national and supranational clinical datasets using i) severity-association networks and ii) GWAS and/or univariate association. Among 365 WW substitutions across 24 genes, we identified 100 (27.4%) showing significant association to hospitalization (Supplementary Table S9). Substitutions found exclusively in clinical settings were far more likely than WW-only substitutions to appear in severity networks (71/246 versus 6/119; Fisher’s exact OR = 7.64, *P* = 2.05 × 10⁻⁸), and WW-group membership overall was strongly associated with network representation (χ² = 33.3, d.f. = 3, *P* = 2.74 × 10⁻⁷). Consistent with this, only four of the 114 unique WW-only substitutions had any severity-related support, and all four mapped to the Spike RBD (S:G339H, S:V445P, S:F486P and S:V445H). In the broader evidence matrix, the strongest marginal association overall was S:Q493E (|log2OR| = 0.944), showing that the full dataset also contains strong single-layer signals detailed in Supplementary Table S9.

We identified 85 unique Spike positions in WW, 33 of these which were associated with hospitalization status in the national dataset using networks, and 13 of them also using GWAS (Fig 4B). These supported positions were concentrated in the RBD. Several of the 33 substitutions detected were connected to each other in the network association analysis in the national dataset. The strongest Spike-Spike interaction linked the N-terminal domain site S:G252V with the RBD site S:L455F (coefficient = 0.365), followed by the RBD pair S:R408S-S:K417N (0.278). Together, these results indicate that WW detects not only single position associations but interacting sites within the Spike. Across epidemiological scales, the supranational network analysis converged on a small group of non-structural protein substitutions, with nsp1:K47R remaining the strongest shared signal and showing prominent age- and sex-dependent interactions in the supranational model.

**Figure 4.**
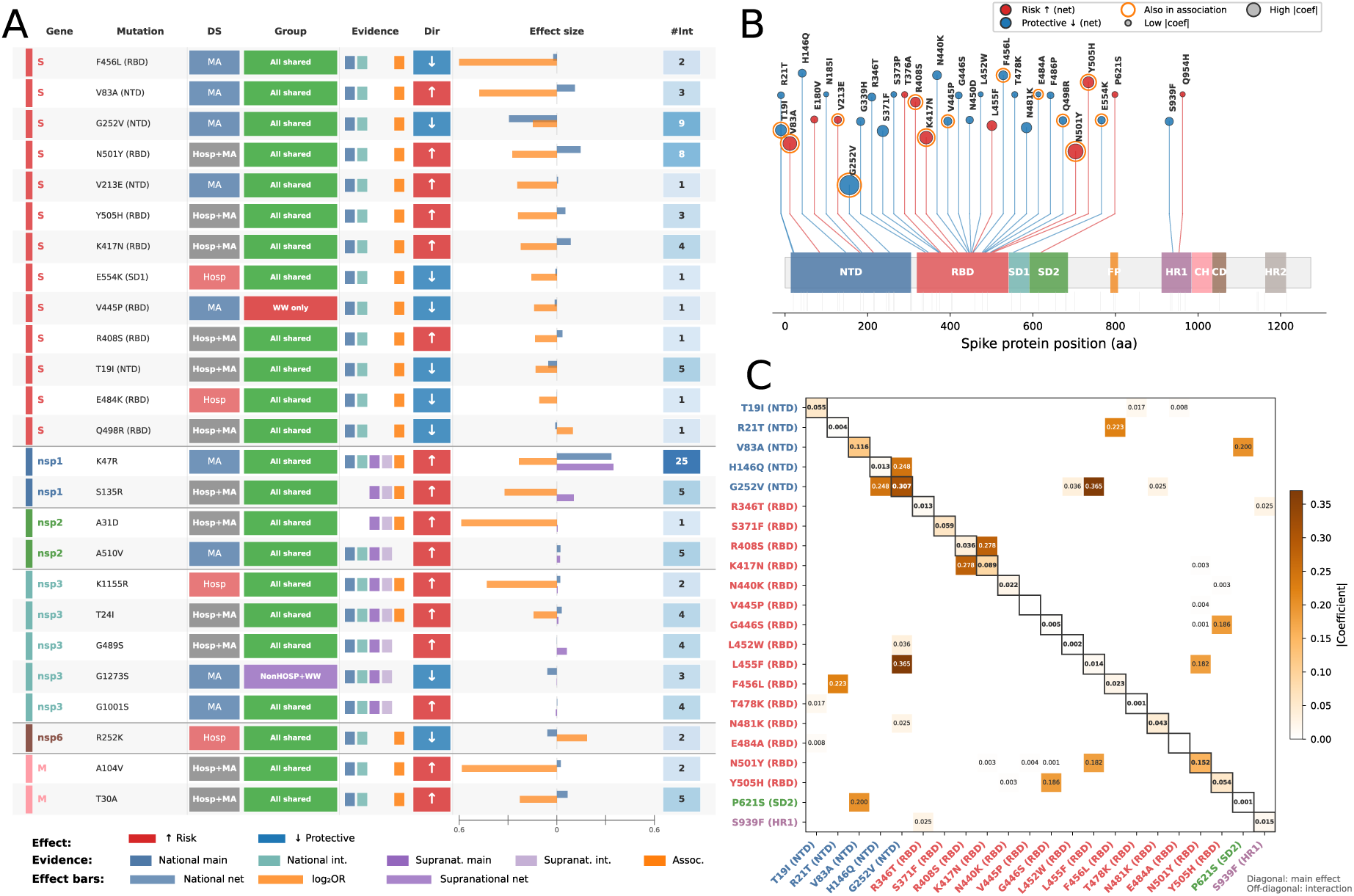
Multi-layer integration of wastewater-detected SARS-CoV-2 mutations linked to hospitalisation-associated severity. **(A)** Compact summary of the 25 detected mutations supported by at least two independent evidence sources across all genes. For each mutation, the panel reports the encoded gene, the amino acid substitution and, for Spike substitutions, the corresponding protein domain; the dataset in which the mutation was detected (MA, metropolitan area dataset; Hosp, hospital dataset); WW group membership; and support across five evidence layers, namely national network main effect (NM), national network interaction (NI), supranational network main effect (SM), supranational network interaction (SI) and univariate association analysis (AS). Horizontal bars show effect size and direction for each evidence layer, with rightward bars indicating risk-associated effects and leftward bars indicating protective effects. The int column indicates the number of significant pairwise interactions involving each mutation in the national network. Mutations are ordered by gene and by the number of supporting evidence layers. **(B)** Linear map of the Spike protein showing the 33 WW-detected Spike positions that overlap with national network evidence. Each circle denotes one amino acid substitution positioned according to residue number, with circle size proportional to the absolute coefficient in the national network. Circle colour indicates the prevailing network direction, with red denoting risk-associated effects and blue denoting protective effects. Orange outlines mark substitutions that are also supported by univariate association analysis. Grey ticks below the protein schematic indicate WW-detected Spike positions without national network support. **(C)** Heatmap of absolute coefficients for the 22 Spike positions retained for interaction analysis in the national network, including positions with at least one significant pairwise interaction and positions with strong main effects. Diagonal cells show absolute main-effect coefficients, whereas off-diagonal cells show absolute pairwise interaction coefficients. Colour intensity reflects coefficient magnitude, and tick labels are coloured by Spike domain.

Zooming in into the 100 WW substitutions associated with hospitalization status, 77 were associated in at least one network, 56 were supported by GWAS and/or univariate association (Supplementary Table S9), and 25 were supported by both (Fig. 4). The Spike contributed the largest share of WW-detected amino acid substitutions (13/25, 52.0%), followed by nsp3 (5/25), with the remaining substitutions distributed across nsp1, nsp2, M, and nsp6. Within Spike, 8 of the 13 prioritised substitutions mapped to the RBD, indicating that the signal highlighted in Fig. 4A is concentrated in functionally interpretable sites rather than evenly distributed across the genome. Among these 25 prioritised substitutions, 23 were shared between wastewater and clinical sequences (All-shared class; group column in Fig. 4A), one was WW-only (S:V445P), and one belonged to the non-hospitalised and WW group, showing that the main signal in Fig. 4A largely reflects substitutions detectable in both wastewater and clinical surveillance. Most of the prioritised substitutions (16/25) combined national-network and univariate-association support, whereas only three were supported by all three evidence layers, namely nsp1:K47R, nsp3:T24I and nsp3:K1155R. All three mapped to non-structural proteins, and nsp1:K47R emerged as the dominant signal overall, with the largest main-effect coefficients in both the national and supranational networks (0.350 and 0.363, respectively) and the greatest network connectivity. By contrast, S:V445P was the only prioritised substitution detected exclusively in wastewater, yet it was still supported by both the national network and the univariate association layer, making it the clearest WW-exclusive candidate. Together, these results indicate that the 25 substitutions shown in Fig. 4A represent high-confidence substitutions associated with hospitalization that are recovered from WW, demonstrating that wastewater surveillance recovers a focused subset of hospitalisation-associated substitutions dominated by shared background variation, while still revealing a small number of potentially relevant substitutions that were absent from contemporaneous clinical sequencing.

## Discussion

Our results align with previous work showing that wastewater surveillance can track SARS-CoV-2 circulation^9–11^ and capture the dominant viral backgrounds, anticipate variant replacement and reveal transmission patterns that are completely represented in patient-based sequencing^12–14,29^. Across the two Valencia surveillance systems, wastewater recovered the dominant Omicron background but showed limited sensitivity for rarer variation. This pattern is biologically plausible because wastewater integrates viral shedding from symptomatic, mildly symptomatic and asymptomatic infections into a single composite population signal. Wastewater is therefore best interpreted not as a substitute for patient-based surveillance but as a community-level readout of the most prevalent circulating viral backgrounds^12,14,23,29^.

A key contribution of this study is the catchment-matched design spanning distinct surveillance contexts and analytical scales. By integrating metropolitan wastewater, hospital wastewater and matched clinical genomes from hospitalised and non-hospitalised infections, we show that much of the apparent disagreement between wastewater and clinical sequencing disappears once closely related sublineages are collapsed into parental lineage constellations. In the metropolitan dataset, Jaccard similarity increased from 0.286 to 0.667 after lineage collapsing, indicating that much of the initial discordance reflected sublineage fragmentation and recombinant nomenclature rather than true failure to recover the dominant Omicron background. This interpretation is consistent with the highly dynamic and fine-grained nature of Pango lineage assignment during Omicron circulation, together with the limited ability of mixed-template wastewater sequencing to resolve every descendant designation with clinical precision^30–32^.

The dual metropolitan-hospital framework also shows that wastewater performance is strongly catchment-dependent. In the metropolitan clinical dataset, hospitalised and non-hospitalised cases shared most lineage constellations and differed only modestly in relative abundance, indicating that these outcome groups were not segregated into distinct lineage pools. In this context, metropolitan wastewater provided the more faithful representation of overall lineage composition. By contrast, the hospital system was noisier at the lineage level, which is expected for a smaller and clinically enriched catchment. Even so, it yielded one of the most notable findings of the study: KP.3 was detected in hospital wastewater in January and February 2024, several months before its first appearance in routine metropolitan clinical surveillance in May 2024. Similar early-warning signals have been reported in other wastewater surveillance systems, where variant emergence has occasionally been detected in environmental samples before routine clinical sequencing captured the same lineages^12,14,33–35^.

A second major contribution of the study is that mutation-level benchmarking was more informative than lineage-level comparison when evaluating wastewater against matched clinical data, because wastewater sequencing often resolves recurrent mutations more robustly than exact lineage composition in mixed samples^21,36^. Applying a coverage-aware framework improved the estimated recovery of clinically observed mutations and clarified that wastewater detection depends not only on mutation prevalence but also on local genome recoverability. In both surveillance settings, wastewater showed high positive predictive value but limited sensitivity, with recovery increasing sharply once mutations became common in the clinical population^29^. In practical terms, wastewater preferentially recovers the common circulating mutational backbone while under-sampling the low-frequency tail where new changes first emerge. This prevalence-dependent sensitivity helps explain why wastewater performs well for dominant lineage constellations and recurrent mutations but less well for rare sublineages and newly emerging variation. Similar detection biases have been described in other wastewater sequencing studies, where mixed viral templates, fragmented RNA and uneven genome coverage limit the consistent recovery of low-frequency variants^13,20,21,29,36^. Overall, wastewater was most informative for common and epidemiologically relevant mutations, whereas rarer emerging variation remained only partially sampled.

Beyond descriptive comparisons, the study also links wastewater-detected variation to clinical outcome. By integrating regional, national and supranational GWAS with regularised interaction networks, we were able to ask not only which mutations were detectable in wastewater but which of those mutations belonged to viral backgrounds enriched among hospitalised cases. One important result is that these hospitalisation-associated signals were only partly reproducible across epidemiological scales^37,38^. This argues against a simple universal catalogue of hospitalisation-associated mutations and instead points to substantial dependence on lineage background, host population structure, immune history and calendar time^27,39,40^. Nevertheless, the repeated appearance of receptor-binding-domain substitutions such as D405N, K417N and R408S in the larger datasets is biologically informative^3,41^. These sites are more plausibly interpreted as markers of antigenically successful Omicron backgrounds than as lineage-independent determinants of severe disease. Mutations at these positions have been implicated in antibody escape and antigenic change during Omicron diversification, reflecting adaptation to population immunity rather than direct effects on virulence^3,42,43^.

The interaction networks reinforce this interpretation by highlighting structured dependencies among Spike sites, consistent with coordinated evolution and background-specific effects rather than isolated one-mutation contributions^3,27^. Notably, some of the strongest edges linked N-terminal-domain and receptor-binding-domain sites, consistent with coordinated evolution across Spike domains rather than independent accumulation of substitutions within a single region^44^. Experimental studies of Omicron evolution have shown that many Spike substitutions act in context-dependent combinations that jointly enhance antibody escape and ACE2 binding^45,46^. These network terms are therefore best interpreted as statistical traces of epistasis, lineage background and shared immune selection rather than as direct causal molecular interactions.

Within this framework, one of the most relevant observations is that wastewater did not merely recapitulate the mutational spectrum visible in contemporaneous clinical sequencing. Most prioritised hospitalisation-associated mutations were shared between wastewater and clinical datasets, but a small subset was detected only in wastewater at the time of sampling. The clearest example was V445P, which remained prioritised despite being absent from contemporaneous clinical genomes. This is biologically plausible because residue 445 lies within a recognised therapeutic-antibody escape hotspot in the Spike receptor-binding domain, and substitutions at this position have been associated with escape from LY-CoV1404 (bebtelovimab)-class antibodies, including complete escape by V445P in XBB backgrounds^47–49^. More broadly, this suggests that wastewater can extend the observable mutational landscape beyond routine patient-based surveillance and recover biologically plausible candidates from transmission chains that remain clinically under-sampled, consistent with previous reports of cryptic or wastewater-exclusive SARS-CoV-2 lineages not represented in contemporaneous clinical datasets^12,16,50^.

The signal also extended beyond Spike. The presence of non-Spike substitutions among the prioritised mutations suggests that the wastewater-supported signal may capture broader evolutionary backgrounds accompanying Spike adaptation rather than antigenic change alone. This interpretation is consistent with experimental work showing that although Spike is the major driver of Omicron immune evasion, non-Spike determinants, particularly nsp6, also contribute to the distinctive replication and attenuation phenotypes of Omicron backgrounds^51,52^. These observations do not establish causality for hospitalisation but support the biological plausibility of a mixed Spike and non-Spike signal emerging from wastewater genomic surveillance. The broader point is that adaptive SARS-CoV-2 backgrounds can be shaped by interacting mutations across the genome rather than by Spike substitutions alone^27,53^.

The limited replication of individual hospitalisation-associated substitutions across the regional, national and supranational cohorts likely reflects the phenotype being tested rather than a failure of the analysis^37,38^. Within the metropolitan dataset, hospitalised and non-hospitalised clinical genomes shared most lineage constellations and showed little abundance divergence, arguing against strong lineage-level segregation by outcome. Hospitalisation during the study period was shaped by age, comorbidity, vaccination history, prior infection, admission thresholds and calendar time, all layered on top of strong viral linkage^39,40,54^. Under those conditions, many apparently protective associations are likely to mark later, more immune-evasive backgrounds circulating in more protected populations rather than intrinsically attenuating mutations^39,55^. This interpretation is consistent with the weak cross-cohort reproducibility of the GWAS hits and explains why the most biologically interpretable signals in this study are concentrated in recurrent antigenic and background-dependent sites rather than in a stable list of universal severity determinants.

Overall, our results refine the role of wastewater genomics during prolonged SARS-CoV-2 circulation^19,23^. Wastewater does not recover all lineage diversity equally, and its performance depends strongly on catchment structure^29,33^. However, when interpreted at the level of collapsed lineage constellations, evaluated through coverage-aware mutation benchmarking and integrated with hospitalisation association analyses, it becomes a powerful prioritisation layer. In our dataset, metropolitan wastewater provided the most stable view of dominant community circulation, whereas hospital wastewater yielded earlier warning for at least one emerging lineage, consistent with previous reports of wastewater-based early detection of emerging variants^12,14^. More broadly, wastewater captured a biologically interpretable subset of viral evolution, including dominant lineage backgrounds, recurrent Spike hotspots and a limited number of clinically missed but relevant mutations^16^. In this sense, wastewater is best viewed not as a surrogate clinical cohort but as a complementary surveillance system that helps prioritise which fraction of ongoing SARS-CoV-2 evolution is most important to track in real time^23^

## Methods

### Study design, datasets and ethics

This retrospective study integrated catchment-matched wastewater and clinical SARS-CoV-2 genomic surveillance in Valencia, Spain, during the Omicron diversification period. Two complementary wastewater catchments were analysed: the Pinedo wastewater treatment plant, representing the Valencia metropolitan area, and the sewer collector of Hospital General Universitario de Valencia (HGUV), representing a healthcare-associated catchment. The local study design was used to compare lineage and mutation profiles between wastewater and clinical surveillance within matched catchments and, in parallel, to evaluate whether wastewater-detected mutations were supported by hospitalisation-associated signals in larger clinical cohorts. For the catchment-matched analyses, the metropolitan dataset comprised 1,933 clinical sequences and 17 wastewater samples, whereas the hospital dataset comprised 449 clinical sequences and 16 wastewater samples. Sampling in the metropolitan system was approximately biweekly, whereas hospital sewer sampling was irregular overall but intensified during January-February 2024. Across both systems, 33 wastewater samples were processed (Supplementary Fig. S1; Supplementary Table S10).

For formal wastewater-clinical comparisons, clinical samples were matched to wastewater dates using ±7 days in the metropolitan dataset and ±2 days in the hospital dataset. To minimise biases introduced by variable wastewater sampling density and uneven genome completeness, analyses were restricted to two predefined windows with consistent sampling and adequate recovery: 9 July 2023 to 8 December 2023 for the metropolitan dataset, and 29 December 2023 to 12 February 2024 for the hospital dataset. These matched windows yielded 11 wastewater sampling dates and 744 clinical genomes in the metropolitan comparison (216 hospitalised and 528 non-hospitalised cases), and 11 wastewater sampling dates with 101 clinical genomes in the hospital comparison. The median timing difference between paired wastewater and clinical sampling points was 10 days in the metropolitan dataset and 9 days in the hospital dataset. Together, these matched local analyses comprised 845 clinical genomes and 22 wastewater sequences. All sequenced samples were evaluated for genome coverage across the SARS-CoV-2 reference genome. In the window-based comparisons between wastewater and clinical sequences, only genomic positions covered in the wastewater sample for each time window were considered in the comparison with clinical genomes (Supplementary Fig. S6).

To support hospitalisation analyses beyond the local matched series, three hospitalisation-annotated clinical cohorts were assembled from GISAID records retrieved on 7 May 2025 for samples collected between 7 June 2023 and 31 August 2024 (Supplementary Table S5). After Nextclade quality control, retention of genomes with reported hospitalisation status, and manual curation of age, sex and hospitalisation metadata, the final analytic datasets comprised a Comunitat Valenciana cohort (n = 4,843), a Spain cohort (n = 10,052) and a global cohort (n = 39,099) (Supplementary Table SX).

This study analysed SARS-CoV-2 genomic data and associated metadata, including metadata retrieved from the GISAID EpiCoV database under its terms of use. Clinical information used in the analyses was limited to de-identified, non-directly identifying metadata, and no identifiable patient records were accessed by the investigators.

The study protocol was reviewed and received a favourable opinion/approval from: (1) the Public Health Research Ethics Committee of the Foundation for the Promotion of Health and Biomedical Research in the Valencian Community (Comité Ético Externo del Biobanco para la Investigación Biomédica y en Salud Pública de la Comunidad Valenciana – IBSP-CV); and (2) the Research Ethics Committee with Medicines (Comité de Ética de la Investigación con Medicamentos) of the Hospital Clínico Universitario de Valencia (Valencia, Spain).

### Wastewater processing, sequencing and primary bioinformatics

The wastewater dataset comprised 33 processed samples collected in Valencia between June 2023 and May 2024 from the HGUV sewer collector and the metropolitan wastewater treatment plant (Supplementary Table S10). For process validation, negative controls consisted of diethyl pyrocarbonate-treated water and the positive control consisted of RNA from an HGUV SARS-CoV-2-positive clinical specimen. For each wastewater sample, 1 L of raw wastewater was collected and transported under refrigerated conditions (5 ± 3 °C) to IATA-CSIC. Viral particles were concentrated according to the Spanish Ministry of Health protocol for SARS-CoV-2 wastewater detection, using aluminium chloride flocculation/precipitation followed by sequential centrifugation, and concentrates were recovered in a final volume of 1-2 mL. Viral RNA was extracted, eluted in 50 µL, transported to I2SysBio on dry ice, and stored at −80 °C until downstream processing.

Clinical respiratory specimens were confirmed as SARS-CoV-2 positive by RT-PCR in the corresponding clinical microbiology laboratories. Viral RNA from clinical and wastewater samples was reverse-transcribed and amplified using the ARTIC tiled-amplicon approach with primer scheme version 5.2. Most clinical samples were sequenced using Illumina short-read technology, whereas a subset of clinical samples was sequenced using Oxford Nanopore Technologies with R9.4.1 and R10.4.1 flow cells. Wastewater amplicons were sequenced using Illumina short-read technology. Illumina and ONT data were processed in amplicon mode with nf-core/viralrecon v3.0.0 against the Wuhan-Hu-1 reference genome (NC_045512.2), including read quality control, trimming, reference-based alignment, primer trimming, duplicate marking, and depth calculation. Variants were called with iVar and consensus genomes were generated with low-support positions masked; optional annotation and lineage assignment steps included bcftools, SnpEff, Pangolin and Nextclade. For clinical samples, SNP calling required minimum depth 20×, minimum base quality Q20, at least four supporting reads with at least two per strand, and minimum allele frequency 5%. The same thresholds were applied to wastewater samples except that the minimum base quality threshold was Q15. Site-level wastewater mutation frequencies derived from this workflow were used for downstream lineage deconvolution and mutation analyses.

### Lineage-level comparison between wastewater and clinical datasets

Clinical consensus genomes passing the inclusion criteria were assigned Nextclade clades as the primary lineage category, with the corresponding Pango lineage retained for interpretation and figure annotation, using Nextclade v3.13.1 and Pangolin v4.3.1^56^. Because wastewater samples may contain mixtures of co-circulating lineages, wastewater lineage composition was inferred from site-level mutation frequencies rather than from a single consensus sequence. These frequency tables were analysed using Freyja v 1.5.1^12^. To stabilise comparisons under low coverage and lineage mixtures, wastewater outputs were analysed both as raw deconvolution calls and after collapsing related descendant sublineages and selected recombinants into broader Omicron lineage constellations according to a predefined remapping dictionary detailed in Supplementary Table S3.

Presence-absence overlap between wastewater and clinical surveillance was quantified using the Jaccard index at both clade and remapped-constellation levels. In the metropolitan dataset, overlap was assessed across three compartments (hospitalised, non-hospitalised and wastewater), whereas in the hospital dataset overlap was assessed between hospitalised clinical genomes and wastewater only. For constellation-level summaries, sensitivity and positive predictive value (PPV) were additionally calculated using the clinical dataset as the reference. To test whether lineage composition differed between hospitalised and non-hospitalised cases in the metropolitan dataset, contingency tables of clade-by-outcome counts were analysed with Pearson’s χ² test and effect size was summarised with Cramér’s V; to account for temporal lineage turnover, the comparison was repeated with a Cochran-Mantel-Haenszel-type stratified analysis using epidemiological week as the stratification variable.

For abundance-based comparisons, each wastewater sampling date within the matched windows was paired to contemporaneous clinical genomes from the corresponding comparison interval, and clade or constellation frequencies were converted to relative-abundance vectors. Concordance between wastewater and clinical surveillance was summarised with Jensen-Shannon divergence, Bray-Curtis dissimilarity, Spearman rank correlation and mean absolute error. Temporal support for the matching framework was assessed by permutation, randomly assigning clinical composition vectors to wastewater dates while preserving the observed set of vectors. To quantify timing differences in emergence, we calculated for each lineage observed in both compartments the difference between its first detection date in wastewater and in clinical surveillance; the median delay was tested against zero using a Wilcoxon signed-rank test.

### Mutation-level comparison between wastewater and clinical datasets

Mutation analyses used a coverage-aware framework designed to reduce bias from uneven wastewater genome recovery. Variant calls, defined as nucleotide substitutions and the corresponding amino acid changes relative to Wuhan-Hu-1, were restricted to callable positions under the study-wide calling thresholds. Two analysis modes were applied: a consensus coverage filter, which retained only mutations at genomic positions covered across all WW samples, and a per-sample time-window approach, in which each WW sample defined a temporal window (±7 for metropolitan area and ±2 days for hospital dataset) and was compared against co-occurring clinical sequences using its own per-sample coverage BED file. In the window-based mode, when the same mutation appeared in multiple windows, the most informative classification was kept (shared > WW-only > clinical-only).

Each mutation was classified into one of three categories: shared (detected in both WW and clinical surveillance), WW-only, or clinical-only. For Dataset I1, which included both hospitalised (HOSP) and non-hospitalised (notHOSP) clinical sequences, mutations were further cross-tabulated into a seven-group Venn diagram (WW × HOSP × notHOSP). For the Hospital dataset, a three-group Venn (WW × HOSP) was used. Concordance between WW and clinical surveillance was quantified using the Jaccard index (J = n_shared / (n_shared + n_WW-only + n_clinical-only)), WW sensitivity (Sensitivity = n_shared / (n_shared + n_clinical-only)), and positive predictive value (PPV = n_shared / (n_shared + n_WW-only)). 95% confidence intervals for sensitivity and PPV were computed using Wilson score intervals. Gene-level enrichment across Venn groups was assessed by two-sided Fisher’s exact test on 2 × 2 contingency tables (target group vs. all others × target gene vs. all others), with Benjamini–Hochberg correction applied per Venn group family. Pairwise comparisons of sensitivity and PPV across analysis modes were performed using Fisher’s exact test with Bonferroni correction (2 metrics × n comparisons).

To assess how clinical prevalence influenced wastewater detectability, each mutation was coded as detected or not detected in wastewater within the matched framework, and clinical prevalence was defined as the proportion of contemporaneous clinical genomes carrying that mutation among all callable genomes at that site. Detection probability was modelled separately for the metropolitan and hospital datasets using binomial generalised linear models with logit link: P(detected | x) = 1 / (1 + exp(−(β₀ + β₁x))), where x is the clinical prevalence (%). Model discrimination was summarised by the area under the receiver operating characteristic curve (AUC), and P_50_ was defined as the clinical prevalence at which the fitted wastewater detection probability reached 0.5. Confidence bands for fitted curves and confidence intervals for P_50_ were obtained by 2,000 bootstrap resamples. Temporal dynamics were further assessed by repeating the coverage-aware comparison across successive matched windows and generating rarefaction-like accumulation curves for shared, wastewater-only and clinical-only mutational sites.

Mutations were also mapped to genes using the SARS-CoV-2 reference annotation to derive gene-level counts, within-gene percentages and recovery metrics. Enrichment of shared wastewater-clinical mutations in specific genes was tested with one-sided Fisher’s exact tests with Benjamini-Hochberg correction. Spike substitutions were assigned to canonical structural domains (NTD, RBD, SD1, SD2, FP, HR1, CH, CD and HR2), and global differences in domain distribution across detection categories were assessed with χ² tests followed by one-sided Fisher tests for group-domain enrichment. For the density displays in Fig. 2, mutation counts were additionally normalised by feature length and compared with length-proportional null expectations using one-sided binomial tests with Bonferroni correction. Shared and group-specific Spike substitutions were mapped onto the prefusion Spike trimer structure (PDB:6VXX) in PyMOL v3.1.

### Hospitalisation association analyses

Hospitalisation-associated mutational analyses were performed using a harmonised workflow across the Comunitat Valenciana, Spain and global consensus-genome cohorts. Amino acid substitutions were encoded as a binary presence-absence matrix and tested against a binary phenotype defined as hospitalised versus non-hospitalised infection. Only recurrent substitutions were retained MAF 0.05, and covariates for adjustment included age, sex and location. To account for viral population structure, cohort-specific maximum-likelihood phylogenies were inferred from masked multiple-sequence alignments using IQ-TREE2^57^ with the GTR+F model, the --polytomy option and 1,000 ultrafast bootstrap replicates, after masking problematic genomic sites. The resulting trees were converted into phylogeny-derived structure terms using distree v1.0.0 (https://github.com/PathoGenOmics-Lab/distree) for downstream association models.

Single-variant association testing was performed separately in each cohort. In the Comunitat Valenciana cohort, each recurrent amino acid substitution was tested using covariate-adjusted logistic regression with significance assessed by likelihood-ratio test and effect sizes reported as log2 odds ratios. In the Spain and global cohorts, single-variant analyses were performed with pyseer v1.3.11^58^ in binary-trait GWAS mode while adjusting for covariates and phylogeny-derived structure. Within each cohort, multiple testing was controlled with Bonferroni correction, using α = 0.05/m, where *m* is the number of tested substitutions; a suggestive threshold of *P* = 1 × 10^-3^ was used for visualisation. Cross-cohort consistency was evaluated by testing enrichment of overlap among significant substitutions with Fisher’s exact/hypergeometric tests, effect-direction concordance with one-sided binomial tests, effect-size concordance with Pearson correlation of log_2_(OR) estimates, and heterogeneity with Cochran’s Q. To capture conditional and potentially non-additive effects, regularised interaction models were fitted in the Spain and global cohorts using glinternet, with λ selected by cross-validation. Non-zero main effects and pairwise interactions were summarised as networks, and robustness was assessed by bootstrap stability selection, considering node or edge selection frequencies ≥0.8 as stable.

### Integration of wastewater-detected mutations with clinical severity evidence

All amino acid substitutions detected in the Valencia wastewater systems were integrated into a combined wastewater mutation catalogue, using the metropolitan dataset (MA) and the hospital dataset (Hosp) as separate observation layers. The integration input comprised 424 wastewater mutation observations (254 in MA and 170 in Hosp), which were collapsed to 365 unique substitutions across 24 genes while preserving dataset-specific detection flags. In the metropolitan dataset, mutations were classified as WW only, hospitalised and WW, non hospitalised and WW or All shared according to their overlap with local clinical compartments; Spike substitutions were also annotated by structural domain. Each wastewater-detected mutation was then annotated with external clinical severity evidence from the Spain and global regularised interaction networks and from the univariate hospitalisation association analyses. For each mutation, we recorded network effect direction, main-effect magnitude, number of interaction partners, strongest interaction partners, and the direction and effect size of univariate association.

The summary table in Fig. 4 was generated by merging the wastewater mutation catalogue with these external evidence layers. Mutations were prioritised if they were detected in wastewater and supported by at least two independent evidence layers, counting the Spain network, the global network and the univariate association layer as separate sources of support. For the network layers, a mutation was considered supported if the GLINTERNET model assigned it a non-zero main-effect coefficient (|β| > 0) or included it in at least one pairwise interaction term, using the cross-validated λ_min_ penalty. For the univariate association layer, significance was determined by cohort-specific Bonferroni-corrected thresholds (*P* < 3.1 × 10⁻⁴ for the worldwide cohort, *P* < 3.4 × 10⁻⁴ for Spain, and *P* < 3.5 × 10⁻⁴ for Valencia; α = 0.05). Each mutation was scored by its number of supporting layers (maximum 3), and only those with ≥2 independent sources were retained in the prioritised panel (25 unique mutations). Within this set, three mutations supported by all three layers (nsp1:K47R, nsp3:T24I, nsp3:K1155R) were designated layer 1, while the remaining 22 with dual-layer support were designated layer 2. Enrichment of clinically supported mutations across wastewater categories was assessed using Fisher’s exact and χ² tests, and concordance between network direction and univariate association direction was evaluated on mutations with valid effect directions in both layers. This framework was intended to prioritise wastewater-detected mutations with the strongest combined support from local detection, multivariable network context and independent clinical severity association.

### Statistics and software

All analyses were performed with custom Python and R workflows. Primary wastewater and clinical read processing used nf-core/viralrecon v3.0.0 in amplicon mode, and downstream analyses used standard packages for data processing, modelling and visualisation, including pandas, numpy and matplotlib, together with IQ-TREE2, distree, pyseer, glinternet and PyMOL. Figure generation for lineage and mutation analyses was implemented in Python, structural displays were prepared in PyMOL, and computational analyses were run on the Garnatxa high-performance computing cluster at I2SysBio.

## Data Availability

All data produced in the present study are available upon reasonable request to the authors

## Acknowledgements

We thank the clinical microbiology services and participating hospitals in the Province of Valencia for sample processing. We also thank the personnel involved in wastewater sampling and logistics at the Pinedo wastewater treatment plant and at the Hospital General Universitario de Valencia sewer collector. We are particularly grateful to José Manuel Marín Noguera, RedMIVA, and the DGSP (Dirección General de Salud Pública) for their assistance and support in the exploitation and use of the data. Computational analyses were carried out using the Garnatxa high performance computing cluster at the Institute for Integrative Systems Biology (I2SysBio), a joint research centre of the University of Valencia and the Spanish National Research Council (CSIC).

## Author Contributions

PRR: Formal analysis, Investigation, Methodology, Software, Visualization, and Writing – original draft; ASC: Data curation, Investigation, Methodology, Software, Writing – review & editing; APC: Data curation, Investigation, Methodology, Writing – review & editing; PCJ: Data curation, Investigation, Methodology, Writing – review & editing; LRR: Data curation, Investigation, Methodology, Writing – review & editing; RA: Data curation, Investigation, Methodology, Writing – review & editing; CVM: Data curation, Investigation, Methodology, Writing – review & editing; CGC: Methodology, Resources, Supervision, Writing – review & editing; IC: Methodology, Resources, Supervision, Writing – review & editing; GS: Methodology, Project administration, Resources, Supervision, Writing – review & editing; FGC: Funding acquisition, Methodology, Resources, Supervision, Writing – review & editing and MC: Conceptualization, Investigation, Funding acquisition, Methodology, Project administration, Resources, Supervision, Writing – original draft.

