## Supplementary Material for "Paired wastewater and clinical genomics across metropolitan and hospital catchments reveals SARS-CoV-2 relevant mutations"

#### Supplemental Tables

**Supplementary Table S1. Concordance summary between wastewater and clinical surveillance by catchment and grouping level.** Global agreement between wastewater (WW) and catchment-matched clinical SARS-CoV-2 lineage profiles for the metropolitan wastewater treatment plant (WW-TP) and the hospital sewer collector (WW-Hosp). Results are shown for two grouping levels, Nextclade clades and a coarse lineage constellation level (Nextclade clade plus a representative Pango lineage after collapsing closely related Pango sublineages; mapping rules in Supplementary Table SC), using the indicated matching windows (Matching_window). For each catchment and grouping level, the table provides the number of WW samples and clinical genomes, the number of categories detected in WW and clinical data, and multiple complementary concordance metrics: presence-absence overlap (Shared, Union, Jaccard), distributional similarity (JSD_mean, Bray_Curtis_mean), rank concordance (Spearman_rho and Spearman_p), and absolute abundance disagreement (MAE_mean_pp and MAE_SD_pp, expressed in percentage points). Bootstrap 95% confidence intervals are reported for Jaccard, JSD, Bray-Curtis, and Spearman rho (columns ending in bootstrap_CI95). Sensitivity and positive predictive value (PPV) are also provided to summarise the coverage-precision trade-off.

**Supplementary Table S2. Window-level concordance metrics for each wastewater sampling date and grouping level.** Per-sampling-date concordance values between WW and clinical lineage profiles, evaluated separately for each grouping level (Grouping) and using the corresponding temporal window (Window). For each WW sample (Sample_ID, WW_date), the table reports the matched clinical window bounds (Window_start, Window_end), the number of clinical genomes in that window (N_clinical_genomes), category counts in WW and clinical data (N_categories_WW, N_categories_clinical, N_shared, N_union), and the resulting concordance metrics (Jaccard, JSD, Bray_Curtis, and MAE_pp in percentage points). The WW categories detected on each sampling date are listed in WW_categories_detected. These window-level values underlie the summary statistics in Supplementary Table A and support time-resolved visualisations of concordance across sampling dates.

**Supplementary Table S3. Lineage mapping and collapsing scheme used to define lineage categories and construct coarse lineage constellation labels.** Deterministic rules used to harmonise lineage labels across WW and clinical datasets and to reduce sparsity from low-frequency Pango sublineages. It includes two components: (i) regex-based rules for assigning Pango lineages to broader lineage families (Constellation regex (Pango)), with example lineages and a Priority_order to resolve overlapping matches; and (ii) the Nextclade clade display-label definitions (Nextclade display label) used to represent clade-specific constellation labels by pairing each Nextclade clade with a representative Pango lineage and collapsing closely related Pango sublineages under that representative (examples in Example_lineages). These mapping rules define the lineage labels used in downstream concordance analyses and Supplementary Fig.s.

**Supplementary Table S4. Master site-level matrix of SARS-CoV-2 mutations included in the wastewater–clinical concordance analysis.** Each row corresponds to one evaluable mutational position in the metropolitan (MA) and/or hospital (Hosp) datasets, with genomic annotation when available, and reports per-dataset detection in wastewater and clinical samples, overlap category, Venn-group assignment, and clinical frequency. For shared mutations, the table additionally provides the number of matched windows in which the site was jointly detected, the total number of evaluable windows, and a persistence label, together with summary flags indicating whether the site was shared, WW-only, clinical-only in both datasets, or discordant across datasets. This site-level matrix underlies the mutation-overlap comparisons shown in Fig. 2.

**Supplementary Table 5. Metadata of SARS-CoV-2 genomic sequences used in the genome-wide association study (GWAS) and clinical comparisons of the metropolitan area available in GISAID.** A total of 39,098 sequences collected between December 2019 and August 2024 across 56 countries were included, organized into three nested datasets: Valencia (n=4,844; 1,727 hospitalised, 3,117 non-hospitalised), National (n=5,208; 3,938 hospitalised, 1,270 non-hospitalised), and Supranational (n=29,046; 9,382 hospitalised, 19,664 non-hospitalised). The hospitalisation phenotype (1=hospitalised, 0=non-hospitalised) was used as the binary outcome for the GWAS. For each sequence, the table reports the GISAID virus name, dataset membership, hospitalisation status, country, province, patient age range (5-year bins), sex, Pango lineage, and Nextstrain clade.

**Supplementary Table S6. Full single-variant association results across the regional, national and supranational cohorts.** Complete results of the amino acid substitution association analyses for hospitalisation outcome in the regional, national, and supranational cohorts. Each row corresponds to one substitution tested in one cohort. Columns report the affected gene, variant identifier, cohort, allele frequency, regression coefficient, standard error, effect size as log2(OR), association P values, the cohort-specific Bonferroni threshold, significance status, and effect direction, together with additional model-output fields retained from the association framework where available.

**Supplementary Table S7. Cross-cohort replication and heterogeneity of Bonferroni-significant amino acid substitutions.** Summary of the amino acid substitutions that reached Bonferroni significance in at least one cohort across the regional, national and supranational hospitalisation analyses. Columns report the number and identity of significant cohorts, per-cohort effect direction, log2(OR), and P values, directional concordance across cohorts, and Cochran's Q heterogeneity statistics.

**Supplementary Table S8. GLINTERNET main effects and interaction terms selected in the national and supranational severity models.** Non-zero main effects and pairwise interaction terms retained by the regularised interaction models fitted to the Spain-wide and supranational cohorts. Rows report the cohort/network, effect type, focal feature, partner feature when applicable, gene annotations, interaction class, and coefficient summaries, including the maximum absolute coefficient and the encoded coefficient vector for each selected term. These entries summarise regularisation-selected statistical dependencies associated with hospitalisation outcome.

**Supplementary Table S9. Full evidence-integration matrix for wastewater-detected mutations linked to clinical severity.** Complete matrix of wastewater-detected mutation entries included in the multi-layer prioritisation analysis. Columns report mutation identity, wastewater dataset, epidemiological group, support from national and supranational severity models and univariate hospitalisation association, effect-size metrics, interaction partners, evidence-layer count, and priority tier.

**Supplementary Table S10.** Metadata of SARS-CoV-2 wastewater samples used in this study. A total of 33 samples were collected between June 2023 and May 2024 in Valencia, Spain, from two sources: a hospital wastewater collector (n=16) and a wastewater treatment plant (n=17). For each sample, the table lists the BioSample and SRA accession numbers, collection date, sampling source, and number of sequencing runs. All samples were sequenced as paired-end reads on Illumina; 18 samples were sequenced in two independent runs. Raw sequencing data are available at the NCBI Sequence Read Archive (SRA) under BioProject PRJNA1444730.

### Supplemental Figures

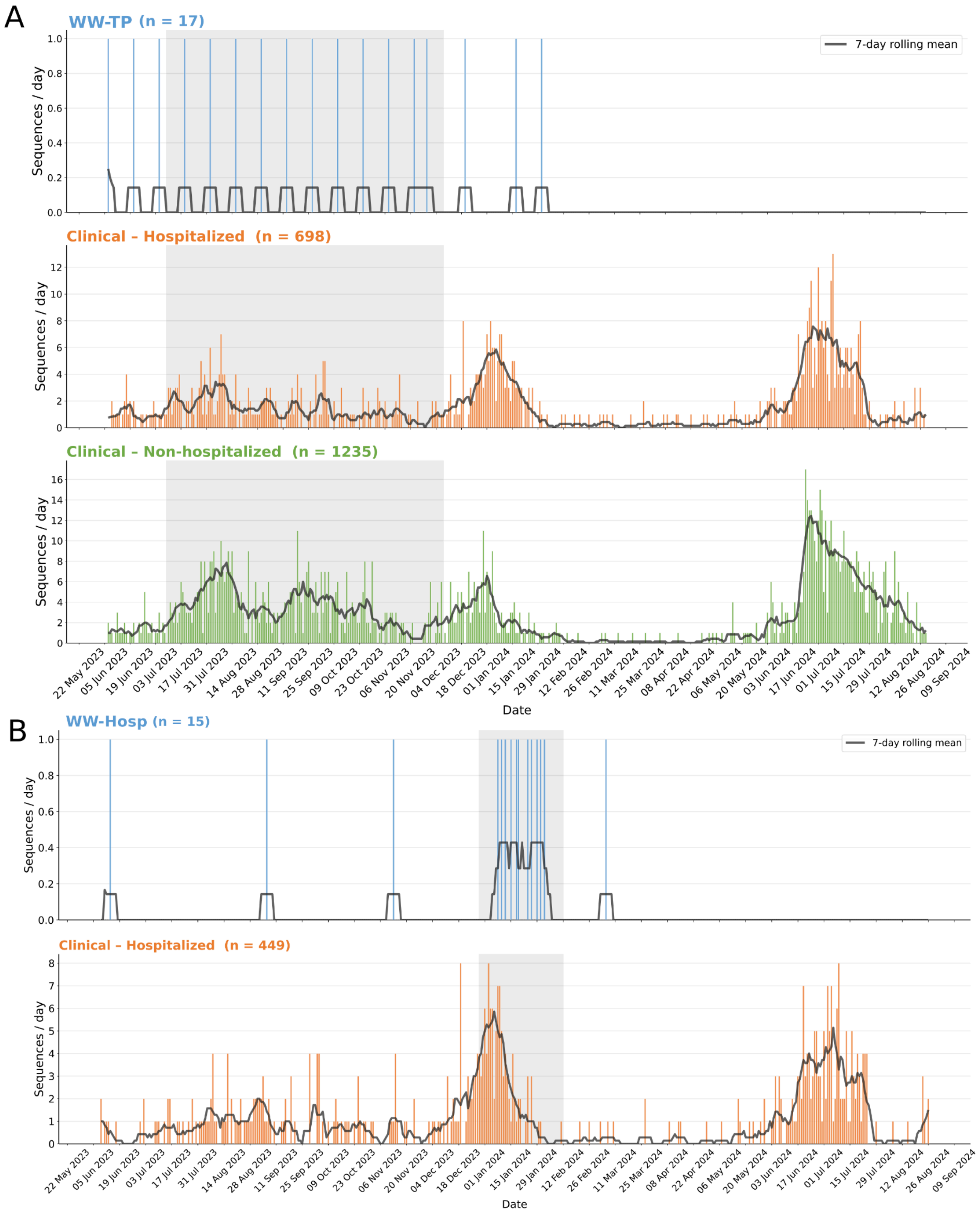


**Supplementary Fig. S1. Temporal distribution of sequenced samples and catchment-matched comparison selected windows.** For the metropolitan area **(A)** and the hospital dataset **(B)**, bars show the number of sequences generated per sampling date in each group. Shaded/zoomed regions indicate the dates included in the formal catchment-matched comparison between clinical and wastewater samples.

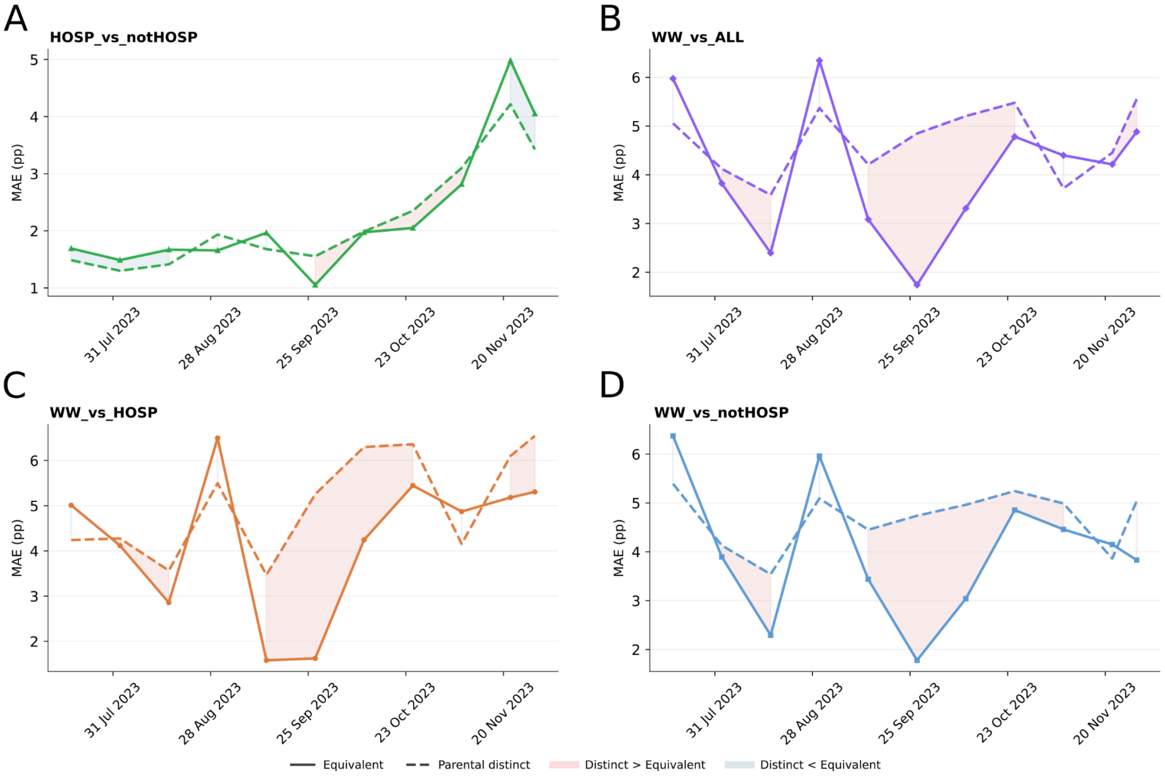


**Supplementary Fig. S2.** Temporal comparison of mean absolute error (MAE) under two parental remapping strategies across four pairwise surveillance comparisons at the metropolitan area from July to November 2023. Panels show hospitalised (HOSP) vs non-hospitalised (notHOSP) **(A)**, WW vs ALL **(B)**, WW vs HOSP **(C)**, and WW vs notHOSP **(D)**. Solid lines indicate the equivalent mode, in which sublineages assigned to the same parental constellation are treated as identical, whereas dashed lines indicate the parental distinct mode, in which each sublineage retains its original identity. MAE is reported in percentage points (pp) and was calculated for each time point. Shaded regions mark intervals in which the parental distinct mode produces higher MAE than the equivalent mode (red) or lower MAE than the equivalent mode (blue). Line colours follow the comparison palette used throughout the study: green for HOSP vs notHOSP, purple for WW vs ALL, orange for WW vs HOSP, and blue for WW vs notHOSP.

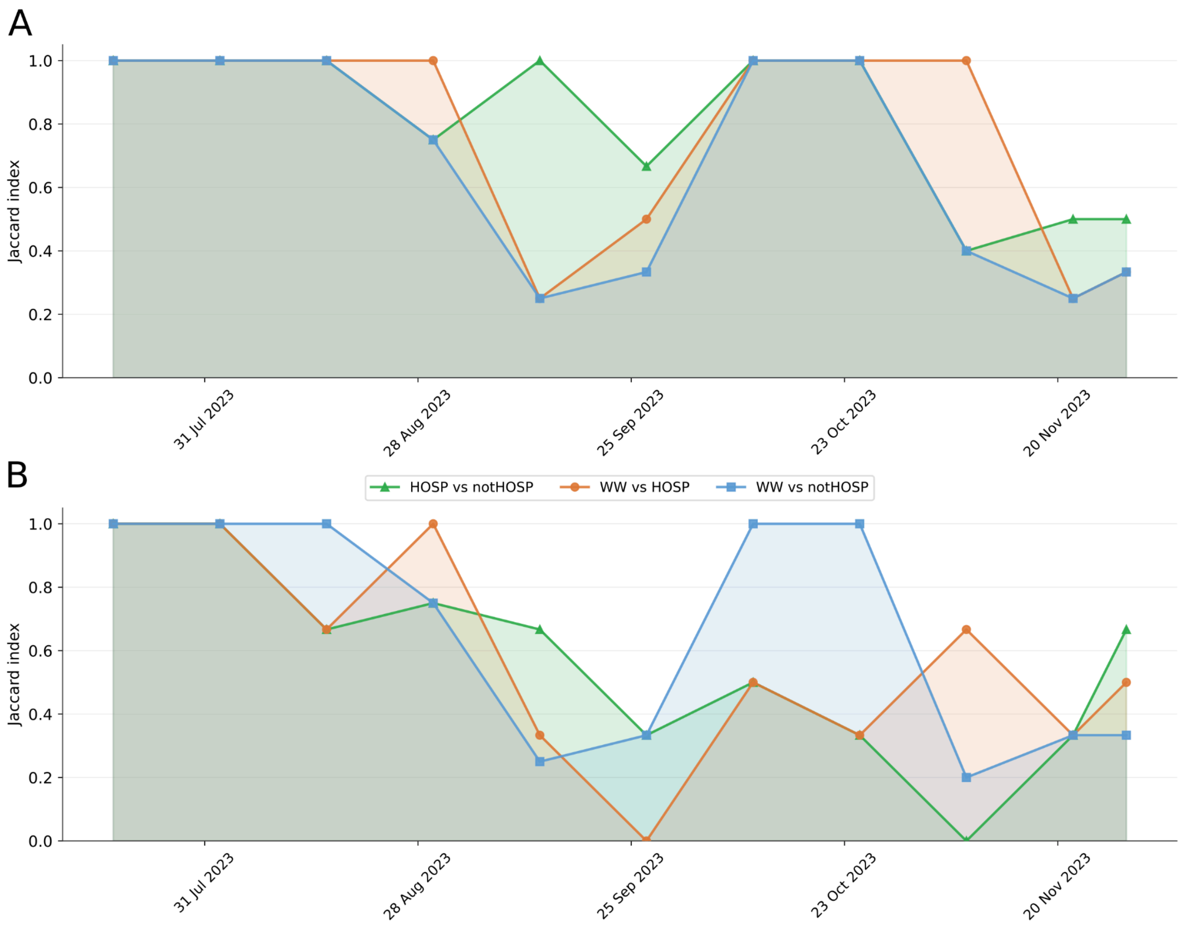


**Supplementary Fig. S3. Temporal variation in Jaccard similarity between wastewater and clinical SARS-CoV-2 lineage profiles in the metropolitan area, July to November 2023.** Each point represents the classic Jaccard index, calculated from presence or absence of lineages with relative abundance ≥0.5% within ±7-day windows centred on each wastewater (WW) sampling date. Three pairwise comparisons are shown: WW versus hospitalised cases (HOSP, orange circles), WW versus non-hospitalised cases (notHOSP, blue squares), and HOSP versus notHOSP (green triangles). Shaded areas under each curve indicate the degree of agreement over time. **(A)** Jaccard similarity after parental lineage reassignment, in which clinical sublineages were collapsed to their corresponding parent constellations to match the resolution of WW-derived lineage assignments. **(B)** Jaccard similarity without parental reassignment, using only directly assigned clinical lineages.

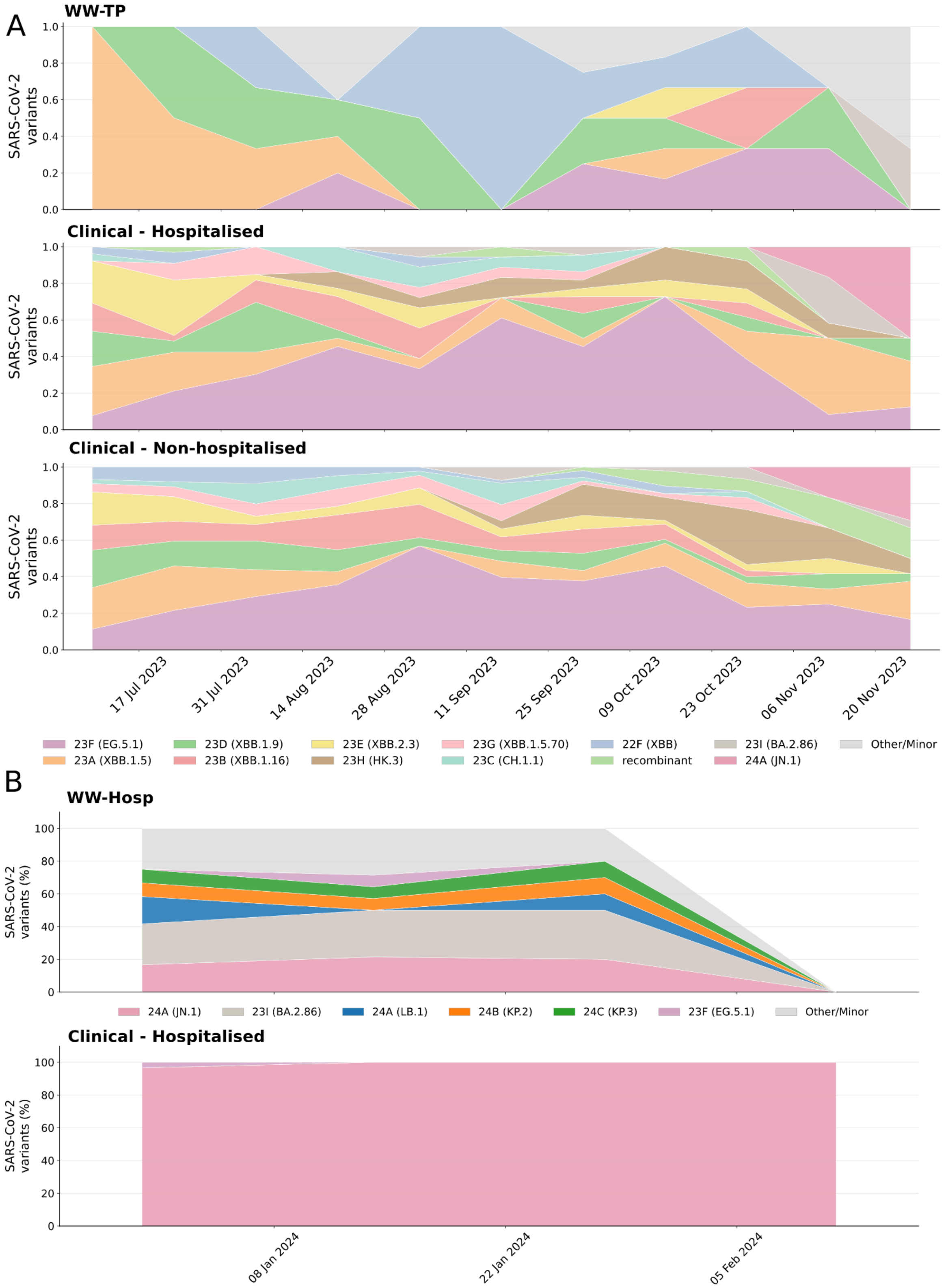


**Supplementary Fig. S4. Temporal dynamics of SARS-CoV-2 clade and lineage-constellation composition across temporally matched sampling windows.**

**(A)** Metropolitan dataset comprising WW_Pinedo wastewater samples and clinical samples from hospitalized and non-hospitalized individuals. **(B)** Hospital dataset comprising ww_general wastewater samples and hospitalized clinical samples. Stacked area plots depict the relative abundance of circulating variant categories over time. Nextclade clades are labeled with representative Pango lineage constellations in parentheses; low-frequency variants are grouped as Other/Minor.

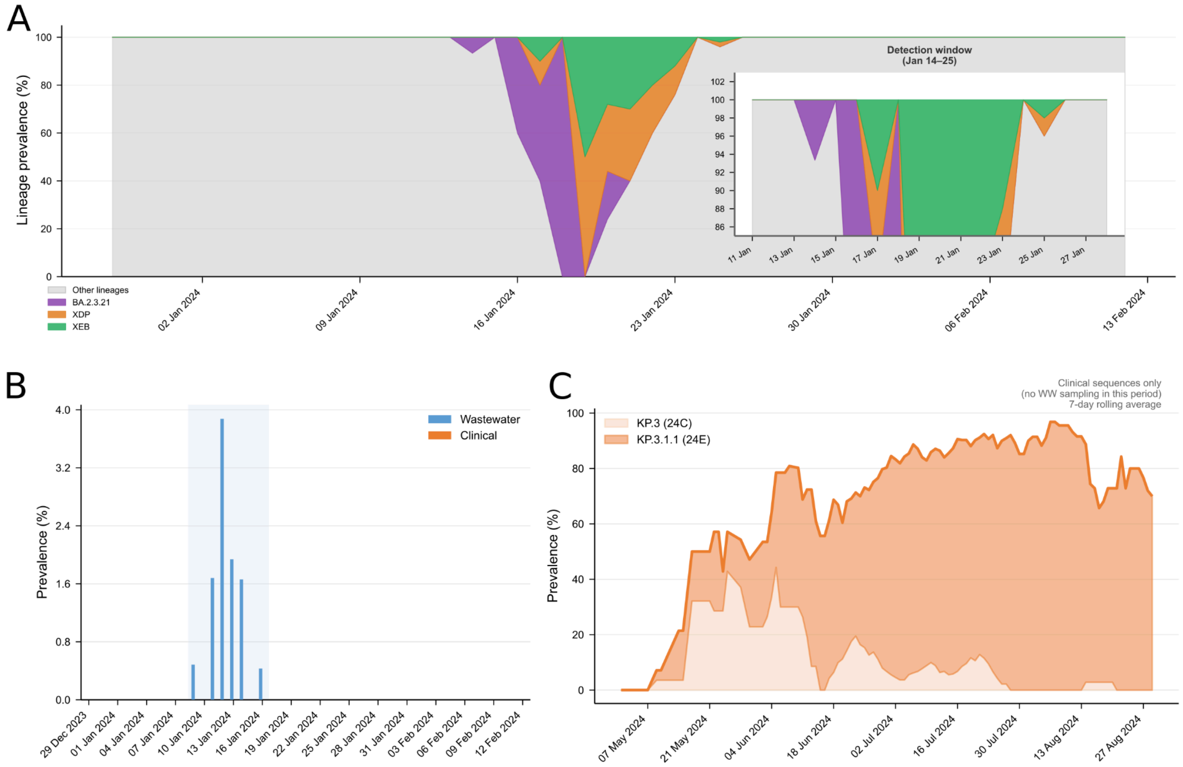


**Supplementary Fig. S5. Wastewater-exclusive SARS-CoV-2 lineage detections in the hospital dataset and subsequent clinical expansion of KP.3-related lineages in the metropolitan area. (A)** Daily lineage composition in wastewater samples collected at the hospital between December 2023 and February 2024. Stacked areas show the relative abundance of SARS-CoV-2 constellations inferred by Freyja deconvolution. Three lineages detected exclusively by wastewater-based surveillance (WBS) and absent from contemporaneous clinical surveillance are highlighted: BA.2.3.21 (purple), XDP (orange), and XEB (green); all remaining lineages are shown in grey. The inset enlarges the detection window from 14 to 25 January 2024, during which these WBS-exclusive lineages collectively accounted for up to ~15% of wastewater abundance while remaining undetected in all sequenced clinical isolates from the same period. **(B)** Daily KP.3 prevalence in the hospital dataset. Bars show KP.3 prevalence in wastewater (blue) and clinical isolates (orange). KP.3 was detected exclusively in wastewater during this interval, reaching a peak prevalence of 3.9%, with no corresponding clinical detections, consistent with an early wastewater signal preceding clinical emergence. The shaded region denotes the WBS-exclusive detection window. **(C)** Temporal dynamics of KP.3-related lineages in metropolitan area clinical sequences from May to August 2024. Filled curves show the 7-day rolling average prevalence of KP.3 (24C, light orange) and KP.3.1.1 (24E, orange). No wastewater sampling was available in the metropolitan area during this period. KP.3.1.1 became dominant by mid-July, reaching >80% prevalence in August 2024, while KP.3 persisted at lower levels.

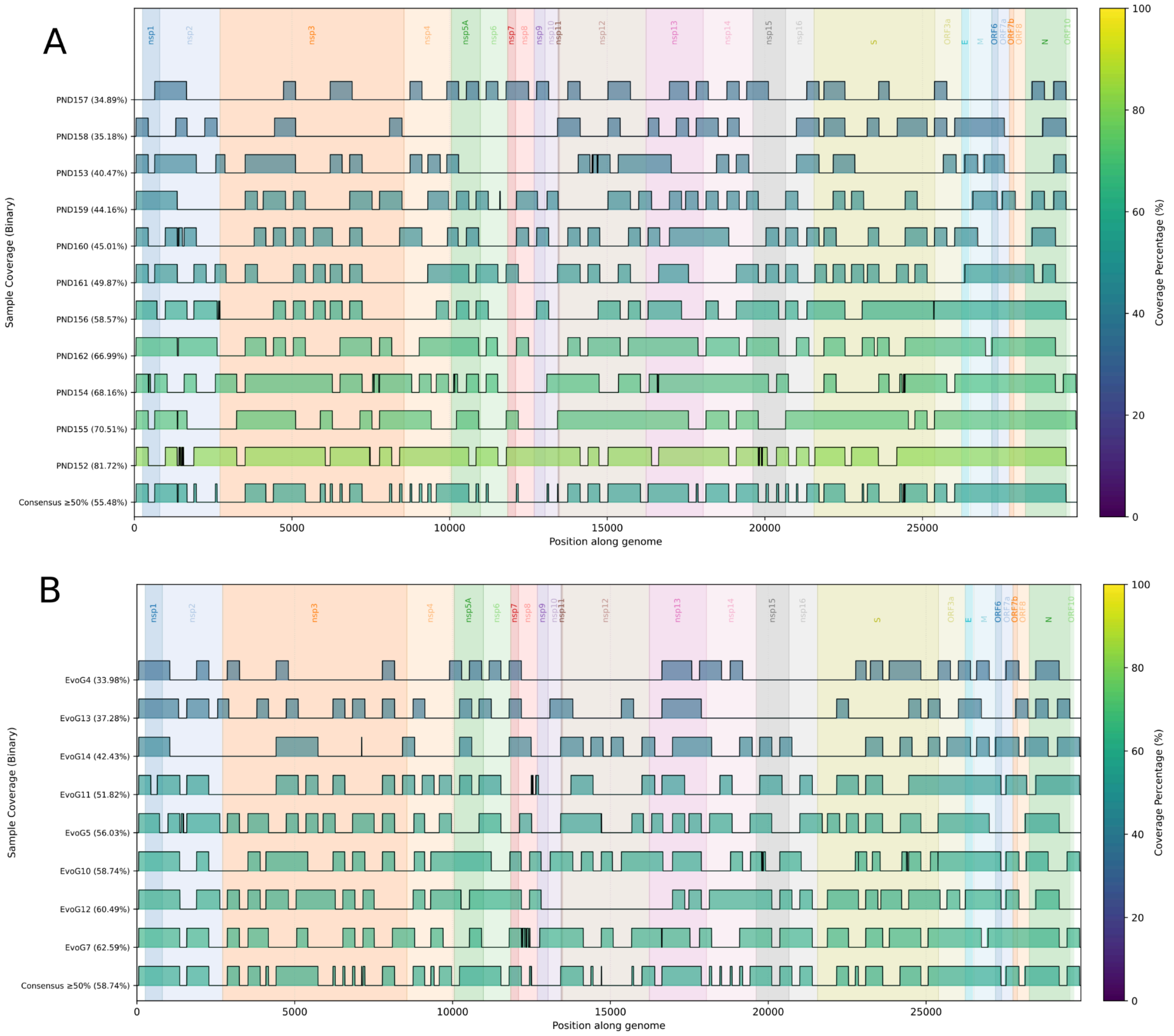


**Supplementary Fig. S6. Genome coverage profiles of wastewater samples included in the WW-TP and hospital collector datasets.** Binary genome coverage along the SARS-CoV-2 reference for each wastewater sample from the Pinedo WW-TP **(A)** and the hospital sewer collector **(B)**. Covered regions (positions with read support) are shown as filled blocks across genomic coordinates; uncovered regions are blank. Gene boundaries are indicated by coloured background shading.

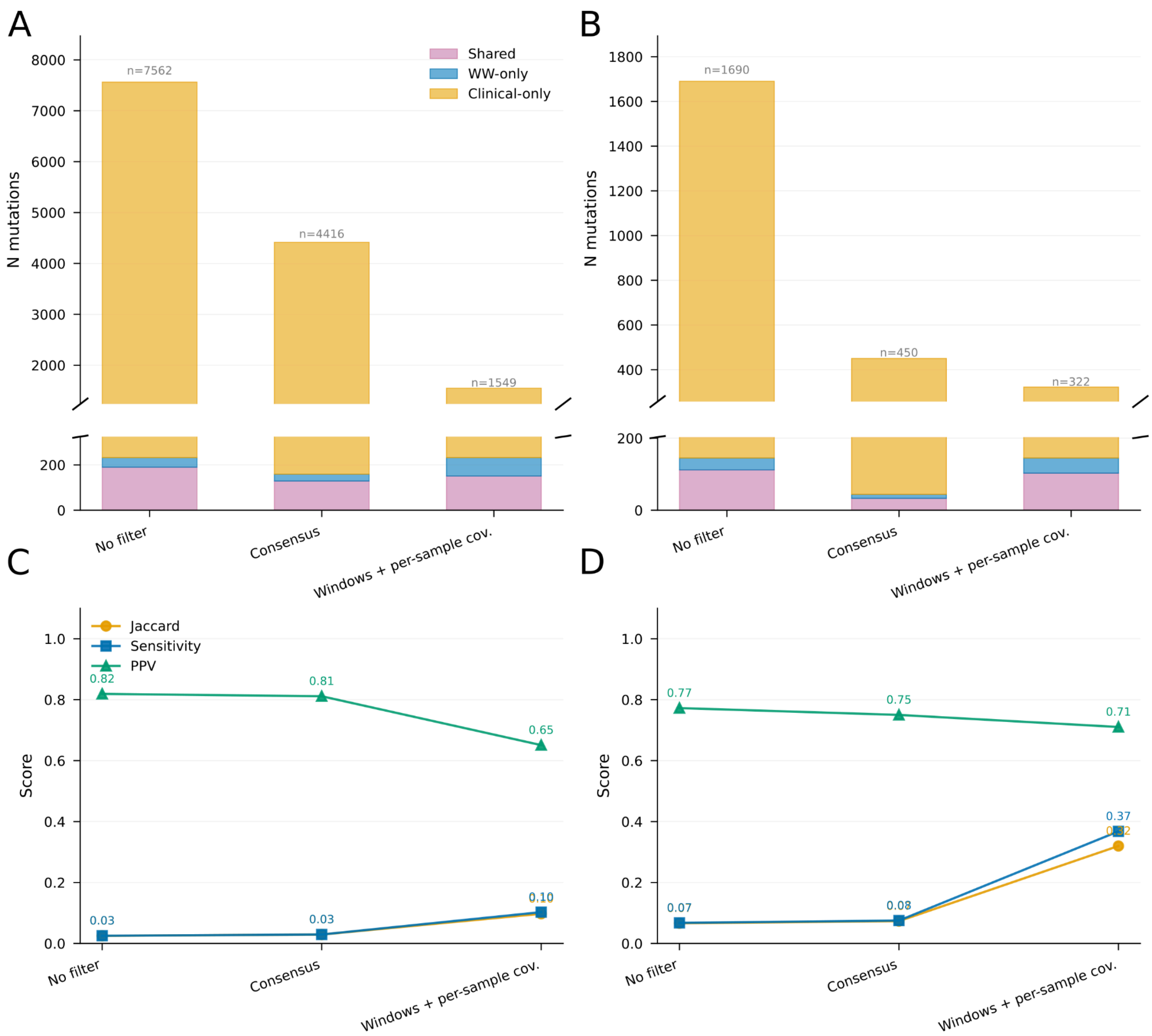


**Supplementary Fig. S7. Comparison of mutation detection and wastewater–clinical concordance across three analytical modes in SARS-CoV-2 genomic surveillance. (A,B)** Stacked bar charts showing the number of mutations classified as Shared, detected in both wastewater (WW) and at least one clinical sample; WW-only, detected exclusively in WW; or Clinical-only, detected in at least one clinical sample but absent from WW, for the metropolitan area **(A)** and the hospital dataset **(B)**. The broken y-axis highlights the relatively small Shared and WW-only fractions, which would otherwise be obscured by the dominant Clinical-only category. Total mutation counts are indicated above each bar. Three analytical modes were compared: No filter, including all mutations regardless of coverage; Consensus, restricting comparisons to genomic positions covered across all WW samples; and Windows + per-sample cov., in which mutations were compared within temporally matched windows using per-sample WW coverage masks (±7 days for the metropolitan area and ±2 days for the hospital). **(C,D)** Concordance metrics for the same analytical modes in the metropolitan area **(C)** and the hospital **(D)**, including the Jaccard index (Shared / [Shared + WW-only + Clinical-only]), Sensitivity (Shared / [Shared + Clinical-only]), and Positive Predictive Value (PPV; Shared / [Shared + WW-only]). The windows-based mode increased Sensitivity and Jaccard concordance relative to the no-filter and consensus approaches while maintaining high PPV, indicating that temporal matching and per-sample coverage masking improve agreement between WW and clinical mutation profiles. The metropolitan-area dataset spans July to December 2023 with biweekly treatment-plant sampling, whereas the hospital dataset spans December 2023 to February 2024 with dense sampling every 2–3 days.

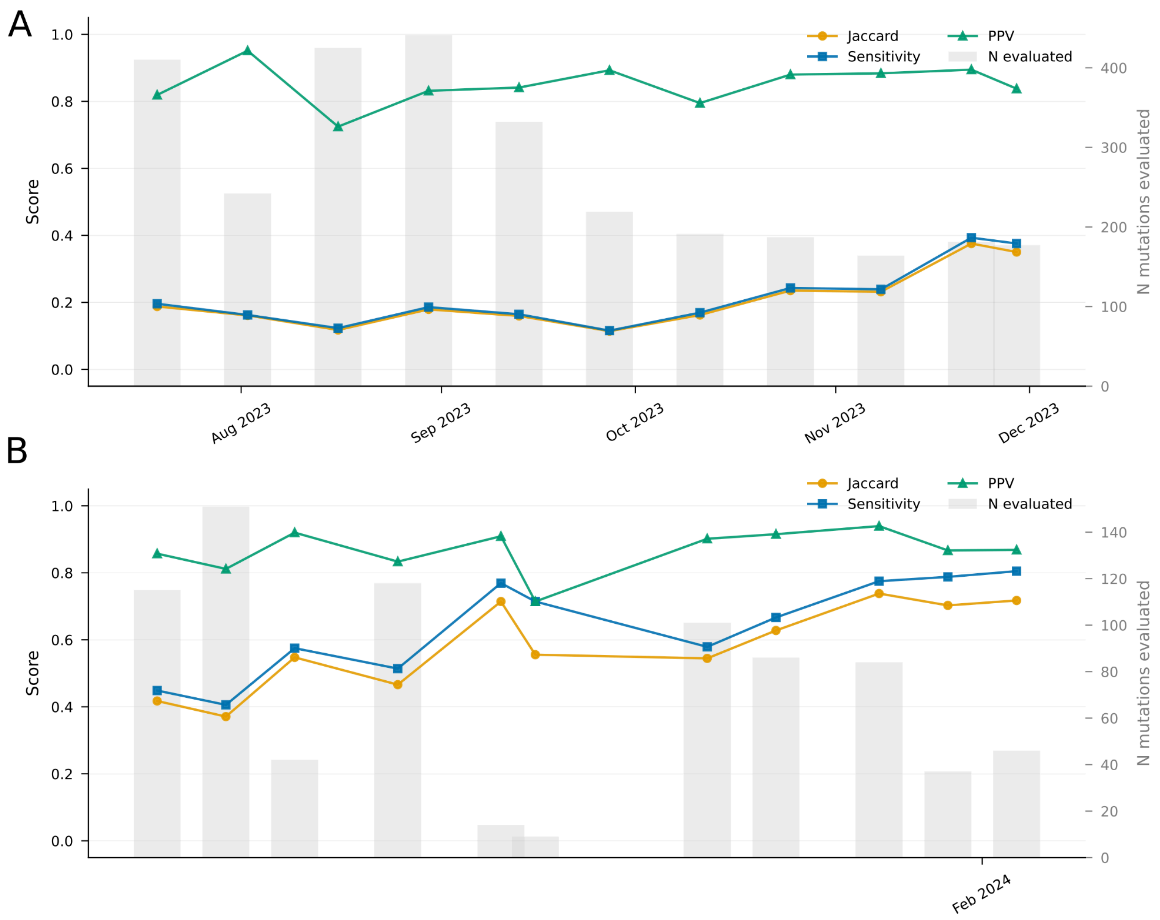


**Supplementary Fig. S8. Temporal dynamics of wastewater-clinical mutation concordance in per-sample time-window analyses.**

**(A)** Metropolitan area, using treatment-plant wastewater sampled biweekly from July to December 2023. **(B)** Hospital, using hospital-sourced wastewater sampled every 2-3 days from December 2023 to February 2024. Shown for each time window are the Jaccard index, sensitivity, and positive predictive value (PPV) between mutations detected in individual wastewater samples and temporally matched clinical samples. Grey bars indicate the number of mutations evaluated per window. Clinical comparisons were restricted to patients sampled within ±7 days in the metropolitan area analysis and ±2 days in the hospital analysis, and to genomic positions covered in the corresponding wastewater sample. PPV remained high across both settings, whereas sensitivity and Jaccard increased over time, consistent with improved concordance as circulating viral diversity narrowed. Concordance was higher overall in the hospital analysis, in line with tighter temporal matching and denser sampling.

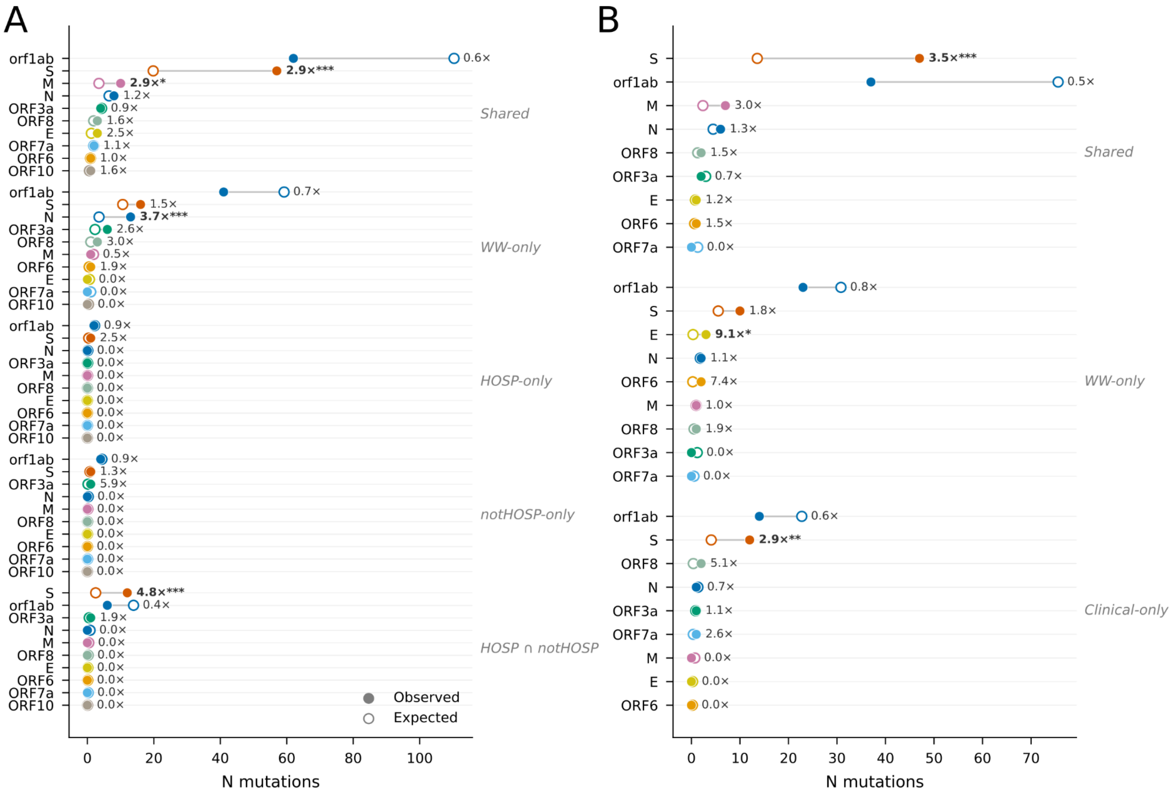


**Supplementary Fig. S9. Gene-level observed versus expected mutation counts across concordance categories.**

Paired dumbbell charts comparing the observed (filled circles) and expected (open circles) numbers of mutations per SARS-CoV-2 gene in the metropolitan area analysis (A) and hospital analysis (B), using the per-sample time-window framework. Expected counts were calculated under a null model in which mutations were distributed in proportion to gene length (bp). Significance of enrichment was assessed using one-sided binomial tests with Bonferroni correction ($P$ < 0.05, P < 0.01, $P$ < 0.001). Fold-enrichment values (observed/expected) are indicated beside each gene. Genes are colour-coded and grouped by concordance category. In the metropolitan area analysis, categories comprise Shared (detected in both wastewater and clinical samples), WW-only (detected only in wastewater), HOSP-only (detected only in hospitalized patients), notHOSP-only (detected only in non-hospitalized patients), and HOSP ∩ notHOSP (clinical mutations detected in both patient groups). In the hospital analysis, which included only hospitalized patients, categories comprise Shared, WW-only, and Clinical-only (detected only in clinical samples). Across both settings, the S gene was enriched among shared mutations, whereas ORF1ab was under-represented relative to its genomic length, consistent with wastewater-clinical concordance being concentrated in genes under stronger selective pressure rather than being uniformly distributed across the viral genome.

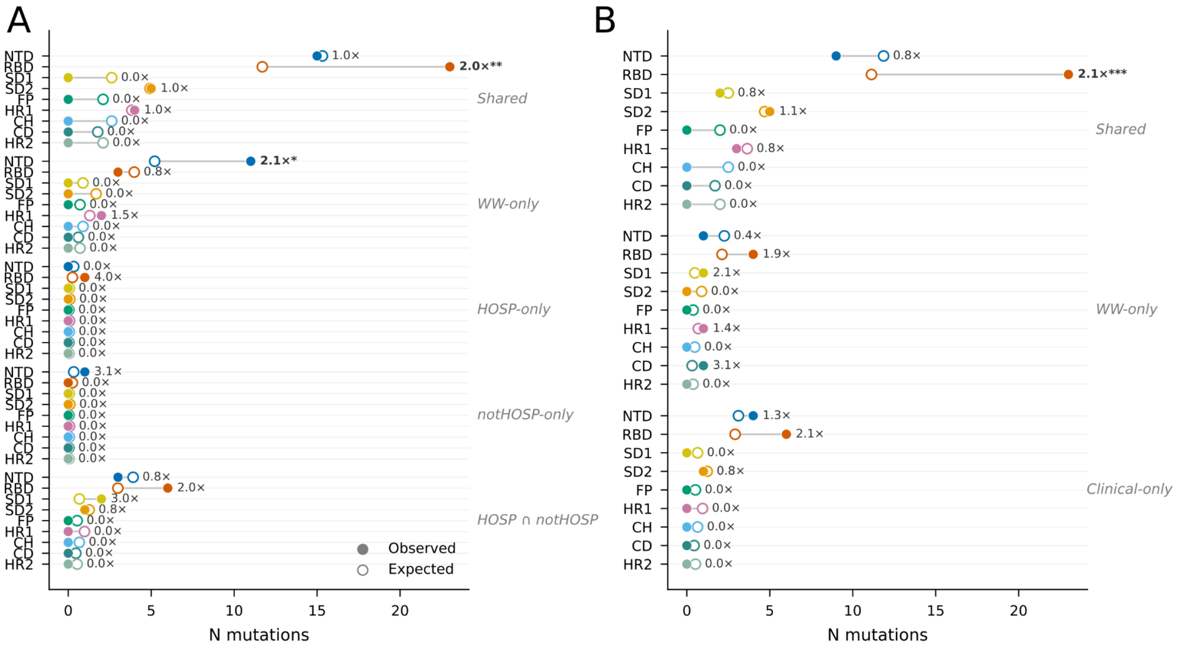


**Supplementary Fig. S10. Spike domain-level observed versus expected mutation counts across concordance categories.**

Paired dumbbell charts comparing the observed (filled circles) and expected (open circles) numbers of mutations across annotated Spike domains in the metropolitan area analysis (A) and hospital analysis (B), using the per-sample time-window framework. Expected counts were calculated under a null model in which mutations were distributed in proportion to domain length (amino acids). Significance of enrichment was assessed using one-sided binomial tests with Bonferroni correction (*P* < 0.05, P < 0.01, *P* < 0.001). Fold-enrichment values (observed/expected) are indicated beside each domain. Domains are colour-coded and grouped by concordance category. In the metropolitan area analysis, categories comprise Shared, WW-only, HOSP-only, notHOSP-only, and HOSP ∩ notHOSP; in the hospital analysis, categories comprise Shared, WW-only, and Clinical-only. Across both settings, the RBD was enriched among shared mutations, whereas most other Spike domains were close to expectation or showed category-specific enrichment, consistent with concordant wastewater-clinical signal being concentrated in functionally and antigenically important regions of Spike. NTD, N-terminal domain; RBD, receptor-binding domain; SD1 and SD2, subdomains 1 and 2; FP, fusion peptide; HR1 and HR2, heptad repeats 1 and 2; CH, central helix; CD, connector domain.

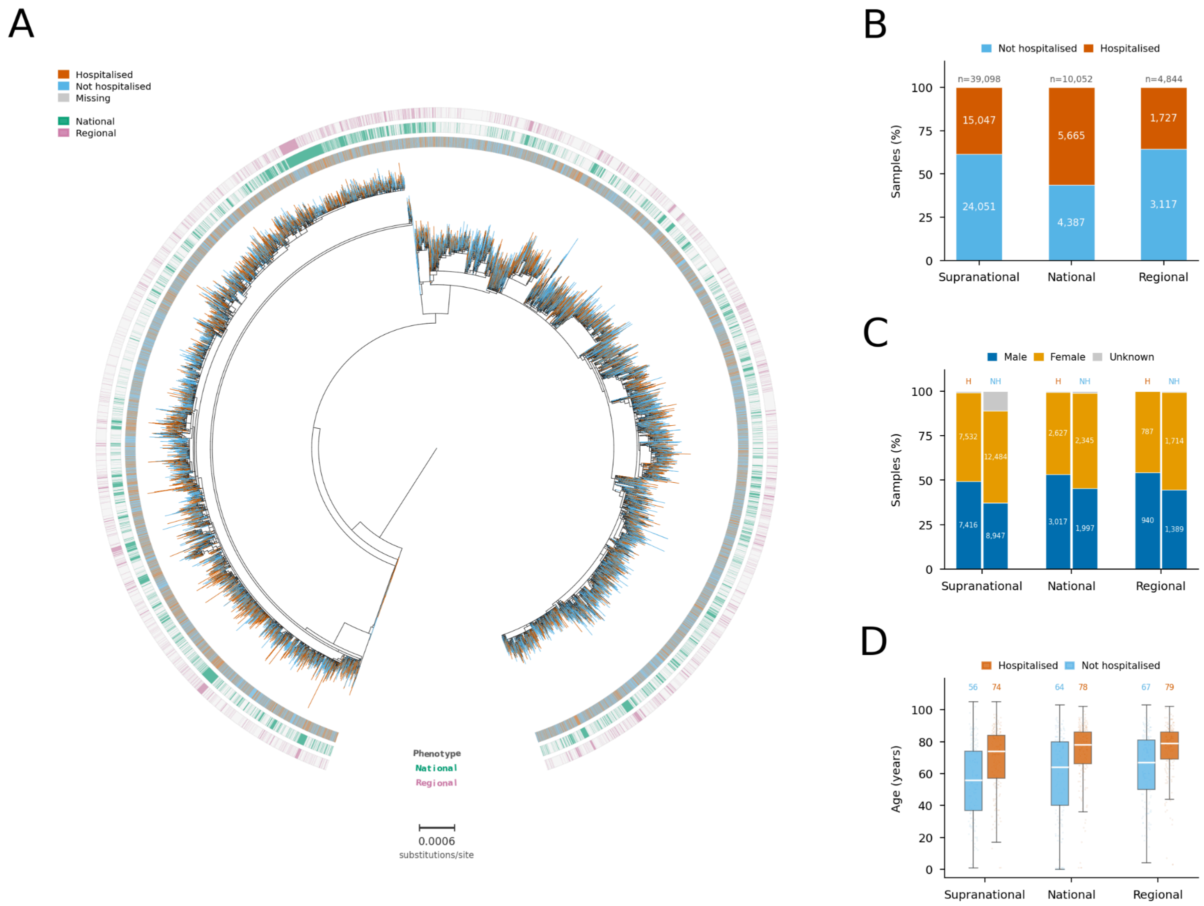


**Supplementary Fig. S11. Phylogenetic and demographic characterisation of SARS-CoV-2 hospitalisation datasets. (A)** Maximum-likelihood phylogeny of 39,099 sequences from the supranational dataset inferred under the GTR+F model. The inner ring indicates hospitalisation phenotype (orange, hospitalised; blue, not hospitalised; grey, missing data). The middle and outer rings denote membership in the national (green; n = 10,052) and regional (pink; n = 4,843) subsets, respectively. **(B)** Proportion of hospitalised (case) and not hospitalised (control) individuals across the three datasets, with absolute counts shown inside bars. **(C)** Sex composition stratified by hospitalisation status (H, hospitalised; NH, not hospitalised) for each dataset. **(D)** Age distribution by hospitalisation status. Box plots show the median (white line), interquartile range, and 1.5× IQR whiskers; coloured numbers above indicate median age (years) per group.

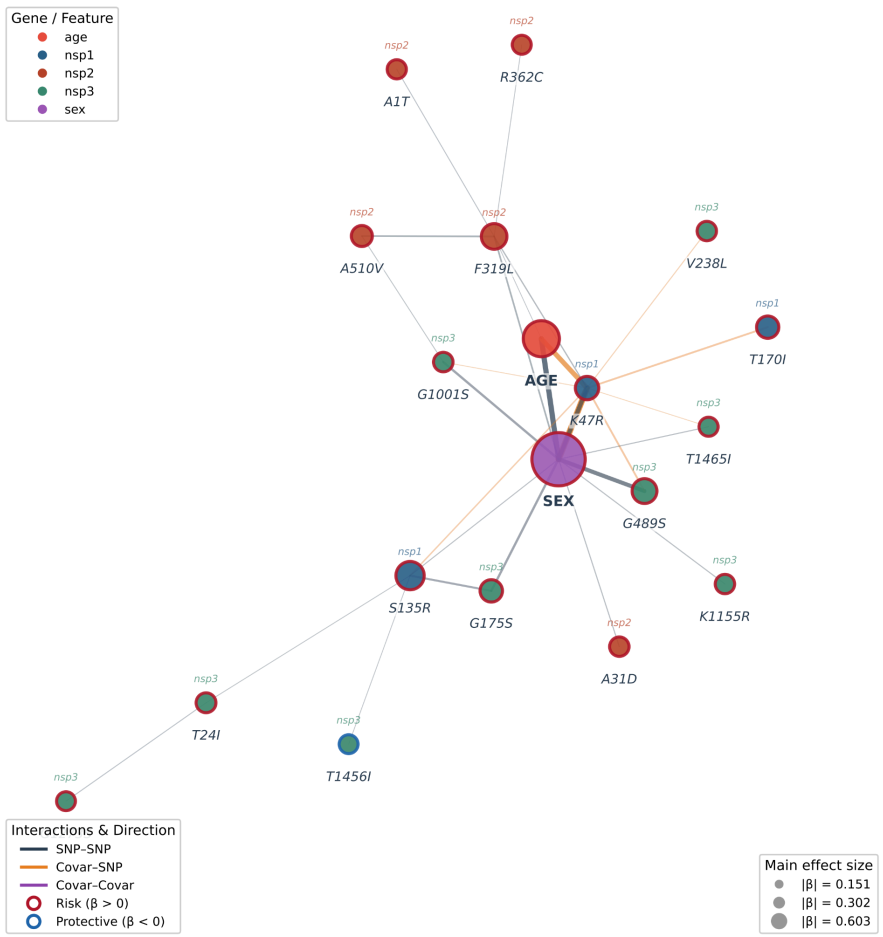


**Supplementary Fig. S12. Interaction network selected by GLINTERNET in the supranational cohort.** Nodes represent predictors retained in the hierarchical penalised model, including amino-acid substitutions and host covariates (age and sex). Node area is proportional to the absolute magnitude of the main-effect coefficient (|β|). Node fill indicates gene or covariate class, whereas node border colour denotes effect direction: red, β > 0, associated with increased odds of hospitalisation; blue, β < 0, associated with decreased odds of hospitalisation. Edges denote predictor pairs with non-zero interaction coefficients selected by the model. Edge width scales with the absolute interaction coefficient, and edge colour indicates interaction class (grey, SNP-SNP; orange, covariate-SNP; purple, covariate-covariate). The network summarises the sparse set of main effects and pairwise interactions associated with the binary hospitalisation outcome in the supranational cohort.
